# Androgen receptor drives polyamine synthesis creating a vulnerability for prostate cancer

**DOI:** 10.1101/2024.12.12.24318845

**Authors:** Rajendra Kumar, Sheila Jonnatan, David E Sanin, Varsha Vakkala, Anoushka Kadam, Shivani Kumar, Susan L Dalrymple, Liang Zhao, Jackson Foley, Cassandra E Holbert, Ashley Nwafor, Srushti Kittane, Elizabeth Penner, Petya Apostolova, Samuel Warner, Chi V Dang, Eneda Toska, Elizabeth A Thompson, John T Isaacs, Angelo M De Marzo, Erika L Pearce, Tracy Murray Stewart, Robert A Casero, Samuel R Denmeade, Laura A Sena

## Abstract

Supraphysiological androgen (SPA) treatment can paradoxically restrict growth of castration-resistant prostate cancer with high androgen receptor (AR) activity, which is the basis for use of Bipolar Androgen Therapy (BAT) for patients with this disease. While androgens are widely appreciated to enhance anabolic metabolism, how SPA-mediated metabolic changes alter prostate cancer progression and therapy response is unknown. Here, we report that SPA markedly increased intracellular and secreted polyamines in prostate cancer models. This occurred through AR binding at enhancer sites upstream of the *ODC1* promoter to increase abundance of ornithine decarboxylase (ODC), a rate-limiting enzyme of polyamine synthesis, and de novo synthesis of polyamines from arginine. SPA-stimulated polyamines enhance prostate cancer fitness, as dCas9-KRAB-mediated inhibition of AR regulation of *ODC1* or direct ODC inhibition by difluoromethylornithine (DFMO) increased efficacy of SPA. Mechanistically, this occurred in part due to increased activity of S-adenosylmethionine decarboxylase 1 (AMD1), which was stimulated both by AR and by loss of negative feedback by polyamines, leading to depletion of its substrate S-adenosylmethionine and global protein methylation. These data provided the rationale for a clinical trial testing the safety and efficacy of BAT in combination with DFMO for patients with metastatic castration-resistant prostate cancer. Pharmacodynamic studies of this drug combination in the first five patients on trial indicated that the drug combination resulted in effective polyamine depletion in plasma. Thus, the AR potently stimulates polyamine synthesis, which constitutes a vulnerability in prostate cancer treated with SPA that can be targeted therapeutically.

## INTRODUCTION

The androgen receptor (AR) is a nuclear hormone receptor that is activated by androgens and exerts downstream effects through its regulation of transcription. It is highly abundant in normal and malignant human prostate, but is also expressed and functional in diverse cell types in men and women. Across cell types, the AR is recognized to drive anabolic metabolism. This function of the AR seems to be largely conserved in prostate cancer (PCa), where it enacts an anabolic transcriptional program and can enhance biosynthesis, such as fatty acid synthesis (*1–3*). Inhibition of the AR by androgen-deprivation therapy not only induces PCa regression, but also results in muscle wasting and metabolic syndrome, highlighting the global role of the AR in regulation of metabolism in humans (*4*).

An interesting aspect of the AR is that it has context-dependent function in PCa. While it enables cancer progression in untreated PCa, we and others have shown that it can paradoxically suppress cancer progression in castration-resistant PCa treated with supraphysiological androgens (SPA) (*5, 6*). This diametric switch in growth regulation appears to occur primarily when ligand-independent AR activity markedly increases as a mechanism of resistance to therapeutic AR blockade (*7, 8*). Clinical trials have demonstrated that periodic administration of SPA (testosterone cypionate 400 mg intramuscularly every 28 days, clinically referred to as bipolar androgen therapy or BAT) results in objective responses for approximately 30% of patients with metastatic castration-resistant PCa and portends a median progression-free survival of 6 months (*9*). BAT is associated with few negative side-effects and several positive side-effects, including improved quality-of-life, and is inexpensive (*9*). Therefore, BAT seems to be an ideal foundation on which to layer additional therapies to enhance anti-tumor effect.

A key effect of SPA associated with therapeutic response is the transcriptional suppression of the protooncogene *MYC* (*8*). MYC, like AR, is widely appreciated to regulate cellular metabolism, driving cell growth and proliferation by enhancing nutrient uptake and amplifying biosynthesis (*10*). Therefore, given that SPA alters two master regulators of metabolism by activating AR and inhibiting MYC, we hypothesized that SPA would result in marked metabolic changes to PCa. This metabolic reprogramming may result in induced metabolic vulnerabilities that could be targeted therapeutically to enhance efficacy of BAT through use of combination therapies.

Here, we report that SPA can alter PCa metabolism in part by increasing de novo polyamine synthesis. Polyamines are small polycationic metabolites that are required for cell proliferation and modulate intra- and intercellular signaling (*11*). We found that SPA increased polyamine synthesis through AR binding upstream of the *ODC1* gene, which encodes ornithine decarboxylase (ODC), a rate-limiting enzyme in this pathway. Although MYC is known to be a positive regulator of ODC, we found that MYC paradoxically suppresses ODC in PCa by antagonizing activity of the AR, which is a stronger regulator of ODC in this context. SPA-stimulated polyamines enhance prostate cancer fitness, as dCas9-KRAB-mediated inhibition of AR regulation of ODC or direct ODC inhibition by difluoromethylornithine (DFMO) enhanced efficacy of SPA. This occurred in part due to increased activity of S-adenosylmethionine decarboxylase 1 (AMD1), which was stimulated both by AR and by loss of negative feedback by polyamines, leading to depletion of its substrate S-adenosylmethionine (SAM) and global protein methylation. These data provided rationale for a clinical trial testing safety and efficacy of BAT in combination with DFMO for patients with metastatic prostate cancer. Pharmacodynamic studies of the first 5 patients on this trial indicated that this drug combination was effective in depletion of polyamines in plasma. Thus, the AR potently stimulates polyamine synthesis, which constitutes a vulnerability in prostate cancer treated with SPA that can be targeted therapeutically.

## RESULTS

### Androgens increase de novo polyamine synthesis and secretion in prostate cancer models

To globally assess how SPA affects the metabolism of PCa, SKCaP-1R patient-derived xenograft tumors of similar size from untreated and SPA-treated mice were subject to global metabolomics by capillary electrophoresis mass spectrometry **(Fig 1A)**. SkCaP-1R is derived from a metastasis of a patient with metastatic castration-resistant PCa who had progressed through multiple systemic therapies including abiraterone, enzalutamide, docetaxel, and carboplatin, grows subcutaneously in castrated NSG mice, exhibits high expression of the AR, and regresses following treatment with SPA (*8, 12*). As anticipated, the metabolome of tumors exposed to SPA was different from that of untreated tumors by principal component analysis **(Fig 1B).** The most increased metabolite by SPA was the polyamine putrescine **(Fig 1C)**. Ornithine, which can be converted to putrescine through ornithine decarboxylase (ODC), was significantly decreased by SPA, as were citrulline and homocitrulline, the products of ornithine transcarbamylase (OTC) downstream of ornithine in the urea cycle **(Fig 1C-D)**. This suggested that SPA may deplete ornithine in favor of production of putrescine rather than citrulline, diverting ornithine to polyamine synthesis rather than progression through the urea cycle.

**Fig. 1.**
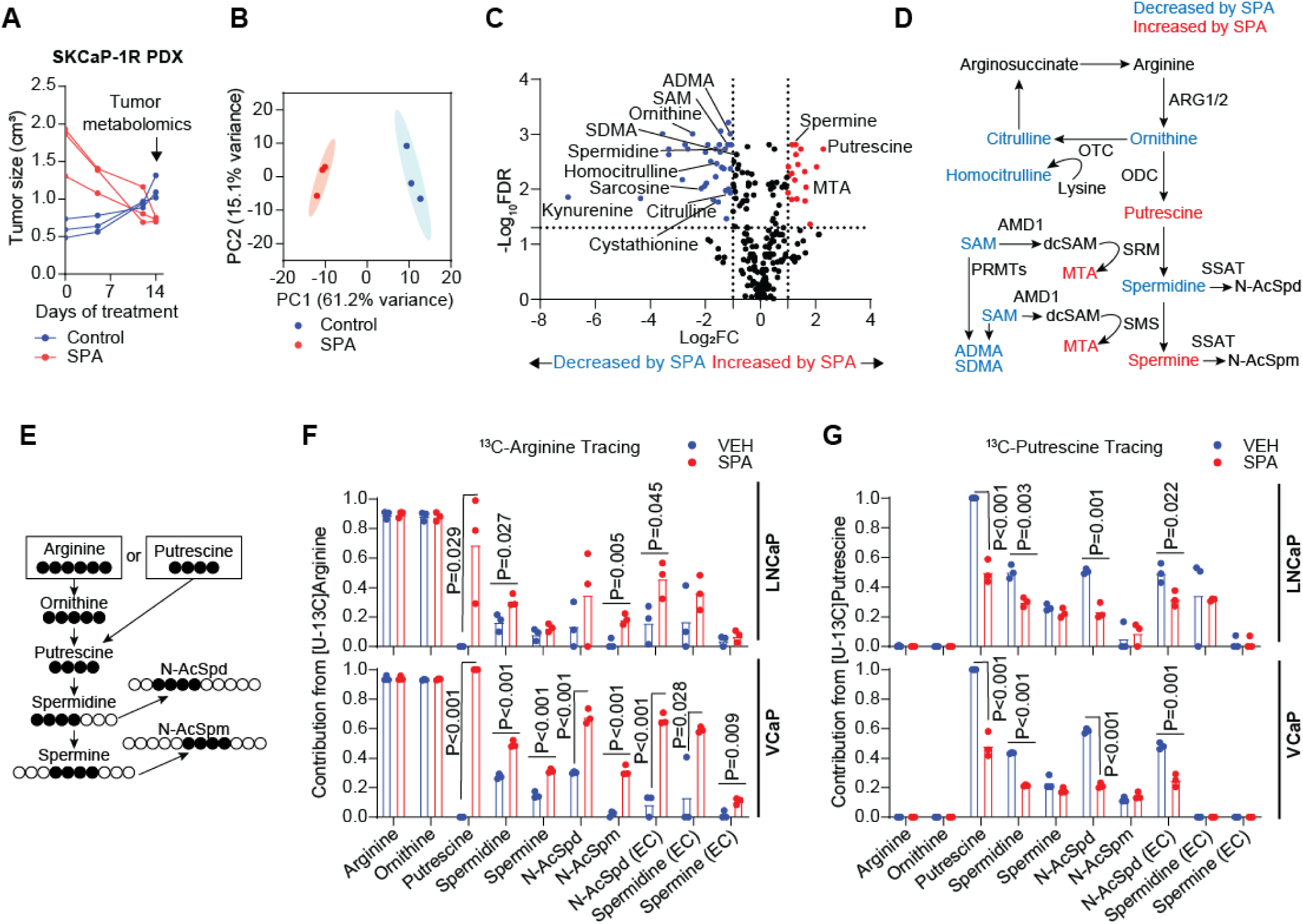
Supraphysiological androgens increase de novo polyamine synthesis in prostate cancer models. A. Tumor size over time of SKCaP-1R patient-derived xenograft tumors growing subcutaneously in the flank of castrated NSG mice untreated (Control) or treated with supraphysiological androgen (SPA; testosterone cypionate subcutaneous pellet). Mice were sacrificed and tumors harvested for global metabolomics following 14 days of treatment. B. Principal component analysis with 95% confidence ellipses of metabolite abundance assessed by capillary electrophoresis mass spectrometry in tumors from (A). C. Volcano plot displaying change in metabolite abundance in SKCaP-1R tumors treated with SPA versus Control from (A). D. Polyamine synthesis pathway schematic displaying metabolites altered in abundance by SPA in tumors from (A). Blue indicates significantly decreased by SPA (Log_2_FC<-1 and - Log_10_FDR>1.3), red indicates significantly increased by SPA (Log_2_FC>1 and −Log_10_FDR>1.3), black indicates not significantly changed by SPA (-Log_2_FC>-1 and <1 or −Log_10_FDR<1.3). E. Isotope tracing schematic. LNCaP or VCaP cells were treated with either vehicle control (VEH; EtOH 0.1%) or SPA (R1881 10nM) in media containing either uniformly-labeled ^13^C-arginine (Silac with 1.1mM U-13C-arginine and 100uM unlabeled putrescine) or ^13^C-putrescine (RPMI, which contains 1.1mM unlabeled arginine, with 100uM U-13C-putrescine). Metabolites were extracted from cells and media at 24 hours and abundance of unlabeled and labeled indicated metabolites determined by liquid-chromatography mass spectrometry. F. Contribution of polyamine synthesis pathway metabolites from arginine in LNCaP and VCaP cells treated with VEH or SPA as per experimental design of (E). N=3 independent experiments. P values by unpaired 2-tailed t test. G. Contribution of polyamine synthesis pathway metabolites from putrescine in LNCaP and VCaP cells treated with VEH or SPA as per experimental design of (E). N=3 independent experiments. P values by unpaired 2-tailed t test.

In the polyamine synthesis pathway, putrescine is converted to spermidine by spermidine synthase (SRM), which is subsequently converted to spermine by spermine synthase (SMS) (**Fig 1D**). Despite increased abundance of putrescine, SPA decreased abundance of spermidine, but increased abundance of spermine, as well as the other product of SMS, methylthioadenosine (MTA) (**Fig 1C-D**). Notably, SAM, the substrate for AMD1 to produce decarboxylated SAM used by SMS to make spermine, was decreased by SPA, as were asymmetric dimethylarginine (ADMA) and symmetric dimethylarginine (SDMA), which are formed by methylation of arginine residues using SAM as a methyl donor **(Fig 1C-D)**. This suggested that SPA may deplete SAM in favor of production of MTA and spermine, rather than methylated proteins, resulting in redistribution of methyl donors in the cell.

In most mammalian cells, ornithine is produced from arginine through arginase 1 and 2, with arginase 2 (ARG2) having much higher expression in the prostate. Thus, to confirm that SPA increases de novo polyamine synthesis in PCa, we traced the fate of exogenously added, uniformly-labeled ^13^C-arginine or ^13^C-putrescine in the AR-positive human PCa cell lines LNCaP and VCaP treated with SPA **(Fig 1E).** Equal labeling of intracellular arginine and ornithine from ^13^C-arginine was observed in vehicle- and SPA-treated cells; however, SPA markedly enhanced labeled putrescine and downstream polyamines from ^13^C-arginine, indicating that SPA increased polyamine synthesis from arginine **(Fig 1F).** In contrast, ^13^C-putrescine tracing revealed that SPA decreased the proportion of intracellular putrescine and downstream polyamines derived from extracellular ^13^C-putrescine uptake **(Fig 1G).** Interestingly, we observed that in these cell lines, SPA not only increased abundance of intracellular polyamines (**Fig S1A)**, but it also increased polyamines in the media of treated cells (**Fig S1B)**, which were also derived from de novo synthesis from arginine (**Fig 1F-G**). Together, these data suggest that SPA increases de novo synthesis and secretion of polyamines from arginine in models of AR-positive PCa to increase abundance of intracellular and extracellular polyamines.

### AR regulates expression of polyamine synthesis enzymes

Over fifty years ago, Pegg and Williams-Ashman demonstrated that androgen deprivation by surgical castration resulted in decreased ODC and AMD1 activity and polyamine abundance in the rat ventral prostate within 6 hours, which was reversible by administration of testosterone to the animals (*13, 14*). Subsequent studies in rodents indicated that androgens can acutely increase *ODC1* mRNA, and prolonged administration of androgens can increase the half-life of ODC protein from 15 minutes to 100-150 minutes (*15–17*). Therefore, we hypothesized that SPA increases de novo synthesis of polyamines in human PCa through AR regulation of enzymes mediating polyamine synthesis, including ODC, AMD1, ARG2, SRM, and SMS **(Fig 2A)**. Indeed, we observed that SPA increased the abundance and activity of ODC over time across several AR-positive human PCa cell lines **(Fig 2B** and **Fig S2A)**. In contrast, SPA increased the abundance and activity of mature AMD1 only in cell lines with high expression of AR, LNCaP and VCaP (**Fig 2B** and **Fig S2B)**. SPA-mediated increases in ODC, AMD1, and polyamine secretion occurred through AR, as they were reduced by inducible knock-down of AR in LNCaP cells (**Fig 2C-D)**. Although ARG2 was not induced by SPA, it appeared to be regulated by AR, as inducible knock-down of AR in LNCaP cells reduced its expression (**Fig 2C),** as did SPA in VCAP cells in association with the auto-downregulation of AR that occurs rapidly in these cells **(Fig 2B)**.

**Fig. 2.**
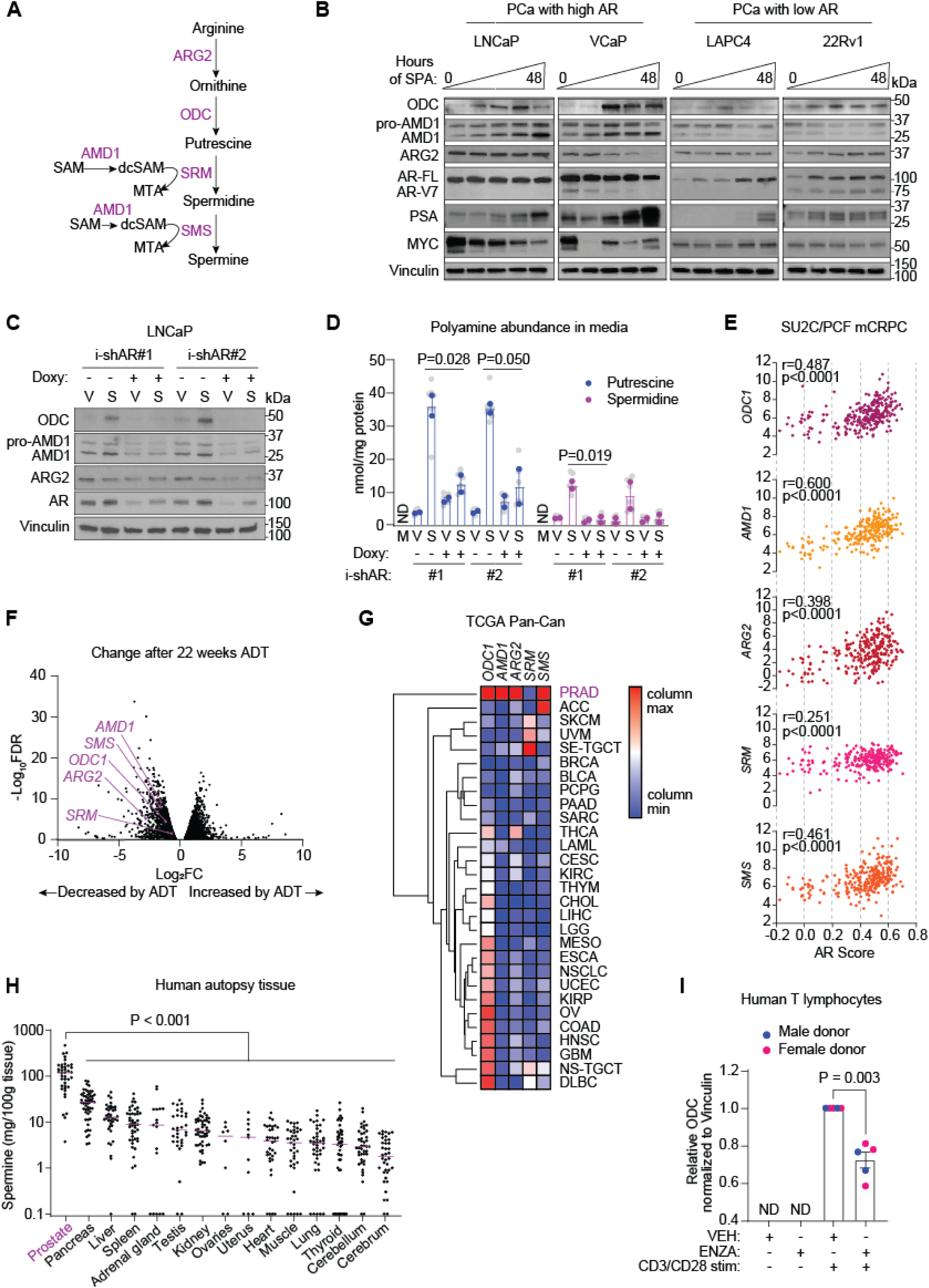
AR regulates expression of polyamine synthetic enzymes. **A.** Schematic highlighting key enzymes that regulate polyamine synthesis in mammalian cells. **B.** Protein expression of ODC, AMD1, ARG2, AR, PSA, and MYC by western blot of prostate cancer cell lines treated with increasing durations of supraphysiological androgen (SPA; R1881 10nM) for 0, 6, 12, 24, and 48 hours. Representative blot of n=3 independent experiments. Vinculin is used as a loading control. **C.** Protein expression of ODC, AMD1, ARG2, and AR by western blot of LNCaP cells expressing doxycycline-inducible shRNA against AR pre-treated with or without doxycycline (doxy) for 48 hours then treated with vehicle control (V; EtOH 0.1%) or SPA (S) for 24 hours. Representative blot of n=2 independent experiments. Vinculin is used as a loading control. **D.** Putrescine and spermidine abundance measured by HPLC in media of cells treated as per (C). M is a cell-free media control. N=2 independent experiments. P values by unpaired 2-tailed t test. Biological replicates indicated in gray with mean of each independent experiment in color. **E.** Correlation of *ODC1, AMD1, ARG2, SRM,* and *SMS* transcript expression (log_2_FPKM) with AR Score in 264 metastatic tumor biopsies of patients with metastatic castration-resistant prostate cancer (mCRPC) in the SU2C/PCF dataset available on cBioPortal. r and p values determined by Pearson’s correlation calculation. **F.** Volcano plot comparing mRNA transcript abundance in tumors of 7 patients with prostate cancer treated for 22 weeks with androgen deprivation therapy (ADT) versus pretreatment samples in the Rajan et al, 2014 dataset. **G.** Heatmap comparing mRNA transcript abundance of *ODC1, AMD1, ARG2, SRM,* and *SMS* across cancer types included in the TCGA Pan-Can dataset available on cBioPortal (n=10,071). Euclidean clustering indicates that prostate adenocarcinoma (PRAD) clusters separately from other cancer types due to its distinctly high expression of these transcripts. **H.** Abundance of spermine purified from human tissues obtained at autopsy of 69 individuals by Hamalainen, 1941. P values by unpaired 2-tailed t test. Quantification from blood, stomach, bone marrow, and thymus are not presented, as fewer than 5 samples were measured. **I.** Quantification of protein expression of ODC by western blot of human T cells cultured for 24 hours with vehicle control (EtOH 0.1%) or enzalutamide (ENZA; 10uM) in the presence of no stimulation or CD3- and CD28-stimulation with dynabeads, displaying data from independent experiments of 5 different donors with indicated sex. P values by unpaired 2-tailed t test.

Next, we assessed whether the abundance of mRNA transcripts of enzymes in this pathway (*ODC1, AMD1, ARG2, SRM,* and *SMS)* seem to be regulated by AR in biopsy samples from patients with PCa. In the SU2C/PCF Dream Team dataset of 264 metastatic biopsies from patients with castration-resistant PCa (*18*), these transcripts positively correlated with an AR activity score (**Fig 2E**). Conversely, in a dataset assessing transcriptional changes to androgen-deprivation therapy in 7 patients with localized PCa (*19*), these transcripts were decreased by androgen-deprivation therapy **(Fig 2F).** Across cancer types in the TCGA Pan-Cancer Atlas study (*20*), PCa exhibited the highest expression of *ODC, AMD1, ARG2,* and *SMS* of any cancer type **(Fig 2G)**, likely due to its uniquely high expression and activity of AR. Notably, in these datasets, *SRM* was not especially abundant in PCa compared to other cancer types, had a relatively weak correlation with AR activity, and had the least change in expression by androgen-deprivation therapy (**Fig 2C-E)**, suggesting that expression of this enzyme is unlikely to be regulated by AR. This may explain why we observed that SPA led to accumulation of putrescine and spermine, but not spermidine, in the SkCaP model (**Fig 1C**). Spermidine enables activation of the translation factor eIF5A, as it is the aminobutyl donor for the hypusination of eIF5A (**Fig S3A)**. In accordance with minimal alteration of SRM expression and spermidine pools by SPA, we noted that SPA also did not alter abundance of hypusinated eIF5A (**Fig S3B**). Altogether these data suggest that AR regulates abundance of polyamine synthetic enzymes, namely ODC, AMD1, ARG2, and SMS, in PCa.

Given that AR regulates polyamine synthesis in the normal prostate of rodents (*13, 14*) and spermine and spermidine were in fact named after their initial discovery in human seminal fluid (*21*), we suspected that regulation of polyamine synthesis is a normal function of the AR that is common between normal and malignant prostate in humans. In 1941, Hamalainen measured spermine concentration in 19 different human tissues obtained during autopsies of 69 individuals with median age of death 40, representation of males and females, with trauma as the leading cause of death (**Fig S4A-C)** (*22*). Statistical analysis of his results confirmed that prostate had the highest abundance of spermine of all measured tissues, with a median concentration of 115.5 mg/100g of prostate tissue, with pancreas ranked as a distant second with a median concentration of 26.7 mg/100g of pancreas tissue (**Fig 2H**). This high abundance of spermine in prostate, which has the highest protein expression of AR across human tissues, is suggestive that regulation of polyamine synthesis is a normal function of the AR that is maintained in cancer.

While AR is most abundant in prostate epithelium, it is also present in diverse cell types with distinct embryological origins such as T lymphocytes (*23*). To assess whether AR regulates expression of polyamine synthetic enzymes in cells beyond prostate epithelium, we isolated human T lymphocytes from peripheral blood of five healthy donors, treated unstimulated or CD3/CD28- stimulated cells with vehicle control or the AR inhibitor enzalutamide for 24 hours, and measured ODC. AR expression was low but detectable in all conditions, however ODC was only detectable in CD3/CD28-stimulated cells **(Fig 2I).** Notably, enzalutamide decreased ODC in CD3/CD28- stimulated cells in all donors **(Fig 2I).** This suggests that AR regulates ODC in T lymphocytes, and the ability of AR to regulate polyamine synthesis is not restricted to cells derived from prostate epithelium, but appears to be a general feature of this transcription factor.

### AR regulates ODC through binding upstream of the ODC promoter

ODC is the first rate-limiting enzyme in polyamine synthesis (*24*), so we sought to define how AR regulates its abundance in PCa. Previous studies identified putative AR response elements (AREs) in close proximity to the transcription start site (TSS) of the *ODC1* gene (*25, 26*). However, AR ChIP-Seq of VCaP cells (*27*) indicated that SPA did not induce AR binding near the *ODC1* TSS **(Fig 3A)**. Instead, SPA induced AR binding at two enhancer sites approximately 4.2 kb (site 1) and 9.8 kb (site 2) upstream of the promoter, which was reduced by concurrent treatment with enzalutamide (**Fig 3A**). These sites were also noted to bind AR in samples from benign (n=7) and malignant (n=13) human prostate (*28*) (**Fig 3A**). To determine whether AR binding at these sites drives increased ODC by SPA, we attempted to block AR binding at these sites by expressing dCas9-KRAB in LNCaP cells targeted to site 1 (sgRNAs A-B) or to site 2 (sgRNAs C-D) **(Fig 3B)**. Basal ODC expression was not altered in these cells, suggesting that dCas9-KRAB did not impede RNA polymerase II binding at the *ODC1* promoter and basal transcription (**Fig 3C**). In contrast, SPA-induced ODC expression was reduced, particularly in cells expressing sgRNAs C-D (**Fig 3D**). Notably, AR ChIP-PCR suggested that SPA-induced AR binding was reduced at both sites upstream of *ODC1* by these sgRNAs (**Fig S5A-B**), suggesting either that dCas9-KRAB can reduce chromatin accessibility across a long range or that there is interdependency of AR binding at these sites. Cells expressing sgRNA-C and sgRNA-D exhibited the greatest reduction of SPA-induced AR binding at both sites, corresponding to the greatest reduction of SPA-induced ODC expression. Cells expressing dCas9-KRAB with a non-targeting sgRNA showed no change in SPA-induced ODC expression (**Fig S5C**). These data indicate that SPA increases ODC expression by increasing AR binding at enhancer sites upstream of *ODC1* to drive its expression.

**Fig. 3.**
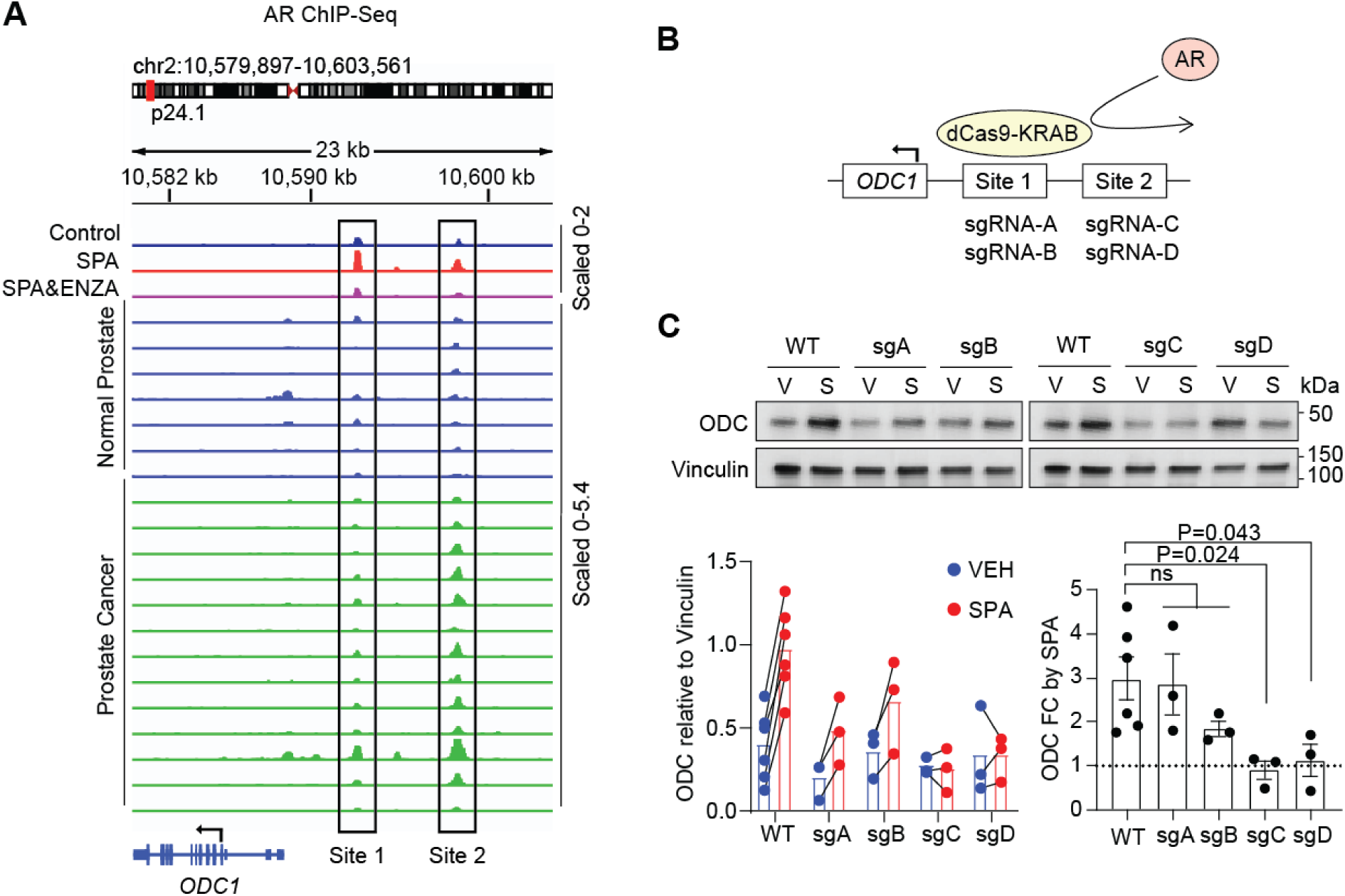
AR regulates ODC through binding upstream of the promoter. **A.** The top 3 tracks display AR ChIP-seq of VCaP cells treated with vehicle control, supraphysiological androgen (SPA; DHT 10nM), or SPA and enzalutamide (ENZA; 10uM) for 12 hours. Data reanalyzed from Asangani et al, 2014. Lower tracks display AR ChIP-seq of 7 histologically normal prostate samples and 13 local prostate cancer samples. Data reanalyzed from Pomerantz et al, 2015. **B.** Schematic depicting dCas9-KRAB-mediated blockade of AR binding. **C.** Protein expression of ODC by western blot of LNCaP cells expressing dCas9-KRAB and indicated sgRNA treated with vehicle control (V) or supraphysiological androgen (S; R1881 10nM) for 24 hours. Vinculin is used as a loading control. Total ODC quantification and ODC fold-change (FC) by SPA of n = 3 independent experiments are displayed below with p values by unpaired 2-tailed t test.

### MYC antagonizes AR-stimulated expression of ODC and AMD1

Historically, MYC, not AR, has been the most widely recognized trans-activator of *ODC1* transcription. MYC binds to a conserved E-box sequence (CACGTG) in the *ODC1* promoter to drive *ODC1* expression (*29–31*). Yet we observed that SPA increased ODC in LNCaP and VCaP cells, despite decreasing MYC (**Fig 2B)**. In VCaP cells (*32*), the downregulation of MYC by SPA expectedly resulted in decreased binding of MYC to the *ODC1* promoter (**Fig 4A**), despite the increase in ODC by SPA in this cell type. Decreasing MYC by siRNA-mediated knockdown in these cells (*32*) did not alter *ODC1* expression, and in fact increased *AMD1* and the canonical AR target *KLK3* (**Fig 4B**). These data indicated that MYC may not be a primary driver of *ODC1* in AR-positive cells. To further probe the role of MYC in the regulation of ODC in the context of SPA, we treated LNCaP cells stably expressing an empty vector (EV) or MYC with vehicle control or SPA and assessed ODC abundance. Forced constitutive expression of MYC actually reduced SPA stimulation of ODC and AMD1 compared with EV control cells, in a similar manner to PSA **(Fig 4C)**. RNAseq of these cells confirmed that *ODC1* and *AMD1* were stimulated by SPA and antagonized by MYC, in a similar manner to *KLK3* **(Fig 4D)**. This surprising reduction of ODC and AMD1 by MYC may be specific to cells that express AR. In these cells, MYC can antagonize the transcriptional activity of AR, possibly through co-factor redistribution when MYC is high, leading to RNA Pol II pausing at promoters of AR-regulated genes (*33, 34*). Indeed, comparison of the fold change of each transcript by SPA in LNCaP-EV versus LNCaP-MYC suggested that the constitutive expression of MYC globally tempered transcriptional changes by SPA (**Fig 4E**). In this comparison, the line of best fit was found to have a shallower slope than the line of unity, with more transcripts showing a smaller change rather than greater change by SPA in cells with constitutive MYC (68% versus 32%) **(Fig 4E)**. This suggests that MYC reduces the transcriptional action of the AR. Therefore, downregulation of MYC by SPA can function as an amplifying circuit to further increase expression of AR target genes, including some genes such as *ODC1* that are typically positively regulated by MYC **(Fig 4F)**.

**Fig. 4.**
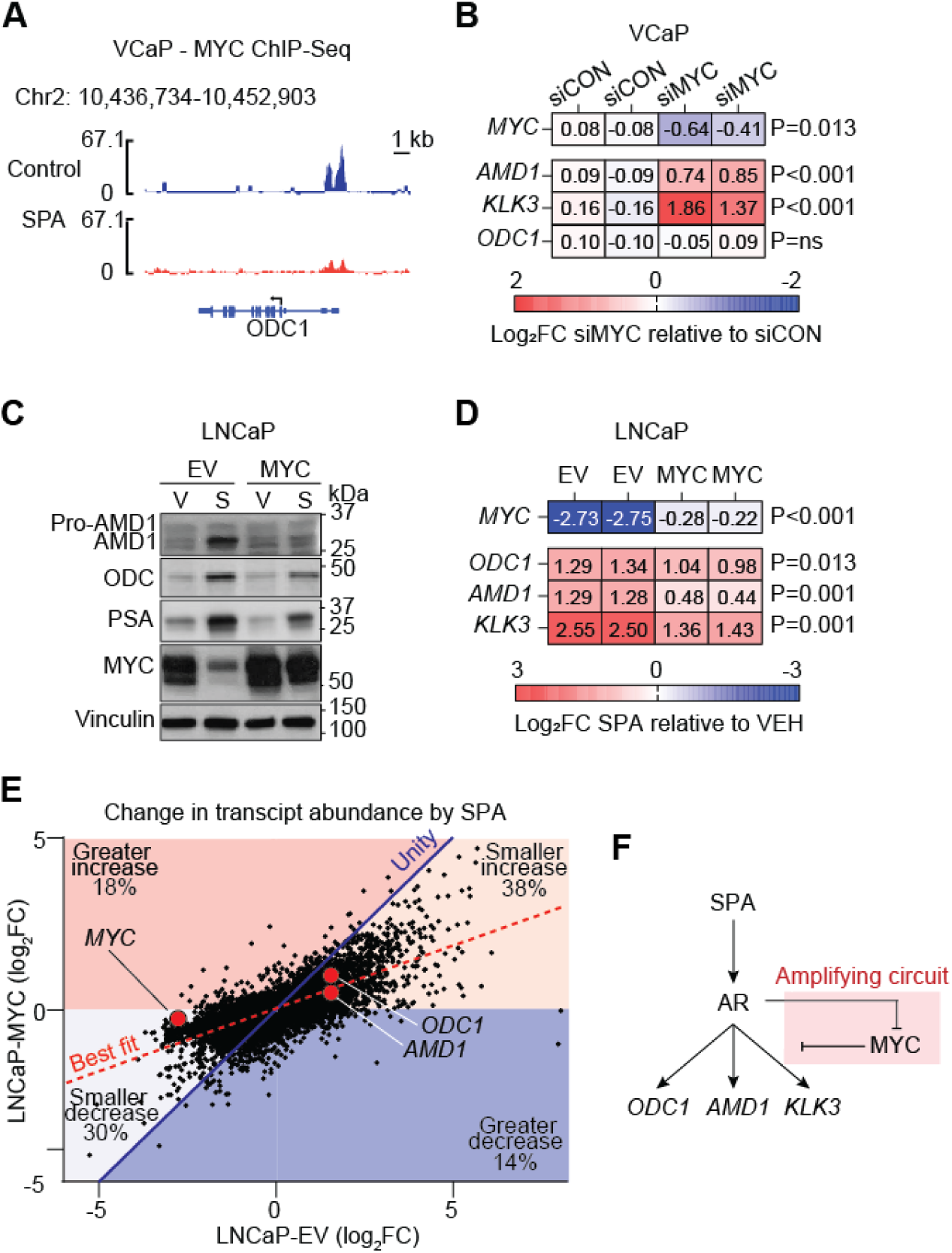
MYC antagonizes AR-stimulated expression of ODC and AMD1. **A.** MYC ChIP-seq in VCaP cells treated with vehicle control or supraphysiological androgen (SPA; DHT 10nM) for 4 hours. Data reanalyzed from Guo et al, 2021. **B.** Change in RNA expression (Log_2_Fold-Change) of *MYC, ODC1, AMD1,* and *KLK3* by RNAseq in VCaP cells with control knock-down or MYC knock-down by siRNA. P values by unpaired 2-tailed t test. Data reanalyzed from Guo et al, 2021. **C**. Protein expression of AMD1, ODC, PSA, and MYC by western blot of LNCaP cells expressing empty vector (EV) or MYC expression vector (MYC) treated with vehicle control (V; EtOH 0.1%) or supraphysiological androgen (S; R1881 10nM) for 96 hours. Vinculin is used as a loading control. Representative blot of n=2 experiments. **D.** Change in RNA expression (Log_2_Fold-Change) of *MYC, ODC1, AMD1,* and *KLK3* by RNAseq in LNCaP cells expressing empty vector (EV) or MYC expression vector (MYC) treated with supraphysiological androgen (S; R1881 10nM) relative to those treated with vehicle control (V; EtOH 0.1%) for 96 hours. P values by unpaired 2-tailed t test. **E.** A comparison of change of individual RNA transcript abundance by SPA in LNCaP-EV cells versus LNCaP-MYC cells. The line of best fit (dotted red) has a shallower slope than the line of unity (solid blue) indicating that many transcripts have reduced change by SPA in LNCaP-MYC cells. Quantitatively, while 18% and 14% of transcripts are more increased or decreased, respectively, by SPA in LNCaP-MYC cells, 38% and 30% of transcripts are less increased or decreased, respectively, by SPA in LNCAP-MYC cells. **F.** Schematic highlighting that the inhibition of MYC by SPA can function as an amplifying circuit to further increase expression of *ODC1*, *AMD1*, and *KLK3* by SPA.

### ODC inhibition enhances PCa growth suppression by SPA

Next, we explored the functional significance of increased ODC and polyamines following SPA treatment. We used LNCaP cells expressing dCas9-KRAB and sgRNAs C or D to block AR binding upstream of *ODC1* as models in which SPA does not increase ODC (**Fig 4C**). We confirmed that ODC and putrescine production was not induced by SPA in these cells even after 6 days of SPA treatment, but could be rescued by concurrent constitutive expression of *ODC1* cDNA (**Fig 5A-B)**. Notably, SPA increased ODC in all cell lines constitutively expressing *ODC1* cDNA, suggesting that the mechanism by which SPA increases ODC is both by increasing *ODC1* transcription and ODC protein stability (**Fig 5A**). Importantly, SPA resulted in greater suppression of proliferation in cells that fail to increase ODC and polyamines (sgRNA-C and sgRNA-D), and this effect could be reversed with constitutive expression of ODC (**Fig 5C**). This indicates that induction of ODC drives relative resistance to SPA. Next, we tested whether pharmacologic inhibition of ODC can phenocopy the effect of blocking ODC induction. Difluoromethylornithine (DFMO) is an analogue of ornithine that is a highly specific irreversible inhibitor of ODC activity (*35*) and is used clinically. Similar to blockade of AR binding upstream of *ODC1*, DFMO enhanced growth inhibition by SPA in LNCaP, VCaP, LAPC4, and 22Rv1 human PCa cell lines (**Fig 5C**). This effect could be rescued by exogenous addition of the ODC product putrescine, indicating that DFMO enhances efficacy of SPA through depletion of putrescine (**Fig 5C**). These data suggest that the induction of ODC and consequent increase in polyamine abundance by SPA facilitates relative resistance to growth inhibition by SPA and SPA can attain greater efficacy when polyamine induction is blocked.

**Fig. 5.**
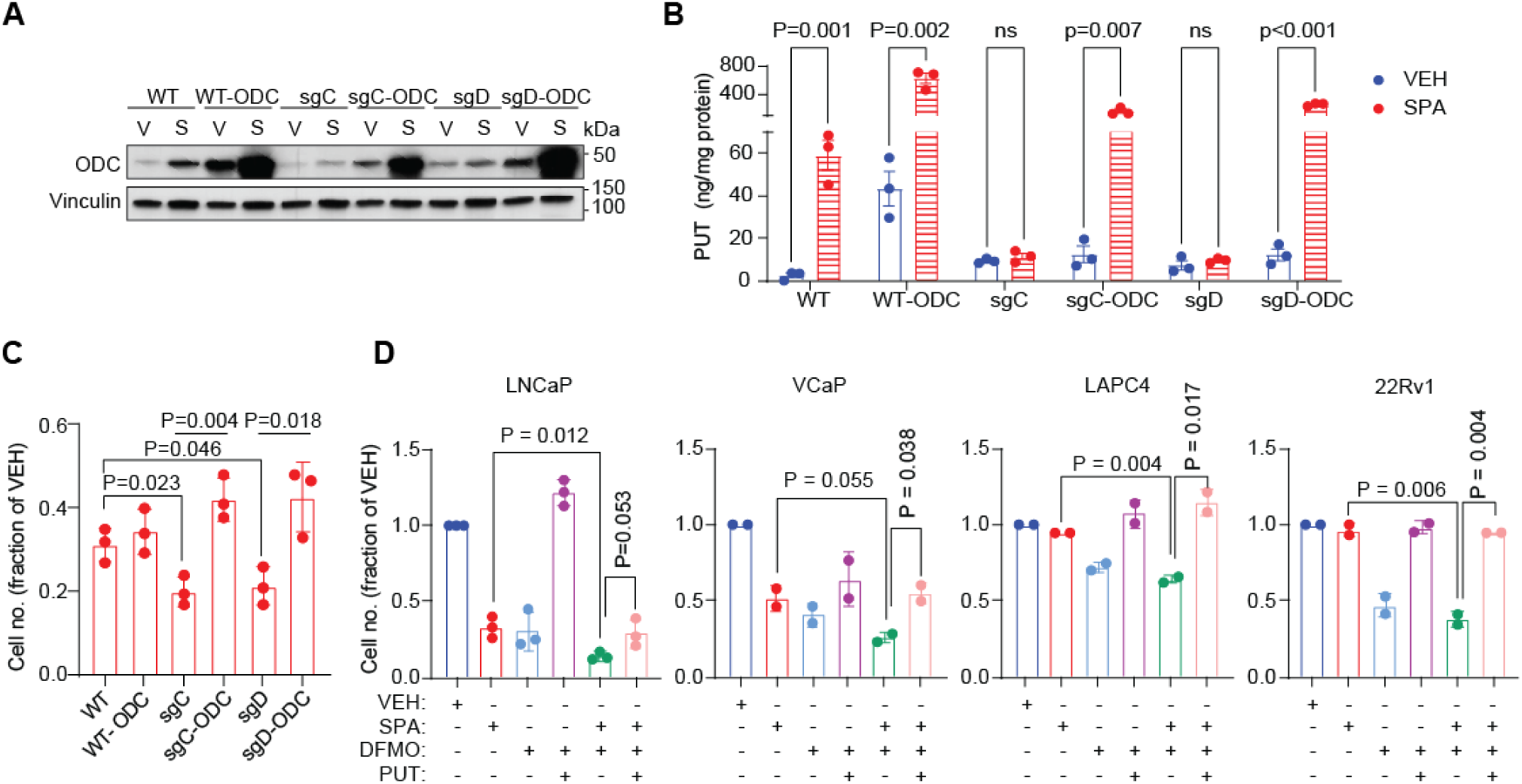
ODC inhibition enhances PCa growth suppression by SPA. **A.** Protein expression of ODC in LNCaP cells expressing dCas9-KRAB and sgRNA-C or sgRNA-D targeting site 2 upstream of *ODC1* with and without concurrent constitutive *ODC1* cDNA expression treated with vehicle control (V; EtOH 0.1%) or supraphysiological androgen (S; R1881 10nM) for 6 days. Vinculin is used as a loading control. Representative blot of n=3 independent experiments. **B.** Putrescine abundance measured by HPLC in media of cells treated as per A. N=3 independent experiments. P values by unpaired 2-tailed t test. **C.** Viable cell number relative to vehicle treatment of cells as per A. N=3 independent experiments. P value by unpaired 2-tailed t test. **C.** Relative viable cell number of indicated cell type treated with indicated combinations of vehicle control (VEH; EtOH 0.1%), supraphysiological androgen (SPA; R1881 10nM), difluoromethylornithine (DFMO 5mM), and putrescine (PUT 100uM) for 7 days. N= 2 or 3 independent experiments. P value by unpaired 2-tailed t test.

### ODC inhibition enhances downregulation of MYC by SPA through depletion of methyl donors

To explore mechanisms by which DFMO enhances growth suppression by SPA, we performed RNAseq of LNCaP cells treated with (1) vehicle control (VEH), (2) SPA, (3) DFMO, (4) SPA and DFMO, (5) DFMO and putrescine, and (6) SPA and DFMO and putrescine for 96 hours. The transcriptome of cells treated with SPA and DFMO, SPA monotherapy, or DFMO monotherapy were different from VEH-treated cells by principal component analysis, with those from cells treated with SPA and DFMO showing the greatest change **(Fig 6A** and **Fig S6A)**. Notably, putrescine supplementation reversed transcriptional effects of DFMO (when used as monotherapy or in combination with SPA) **(Fig 6A)**, indicating that transcriptional changes due to DFMO seem to be on-target effects of ODC inhibition and consequent putrescine depletion. While there was definite overlap of transcripts differentially expressed in cells treated with SPA and DFMO and cells treated with each monotherapy, many transcripts were exclusively altered by combination therapy or monotherapy (**Fig S6A**), suggesting that SPA and DFMO combination therapy has a unique effect on the transcriptome of PCa, that is not simply the summation of effects of each monotherapy.

**Fig. 6.**
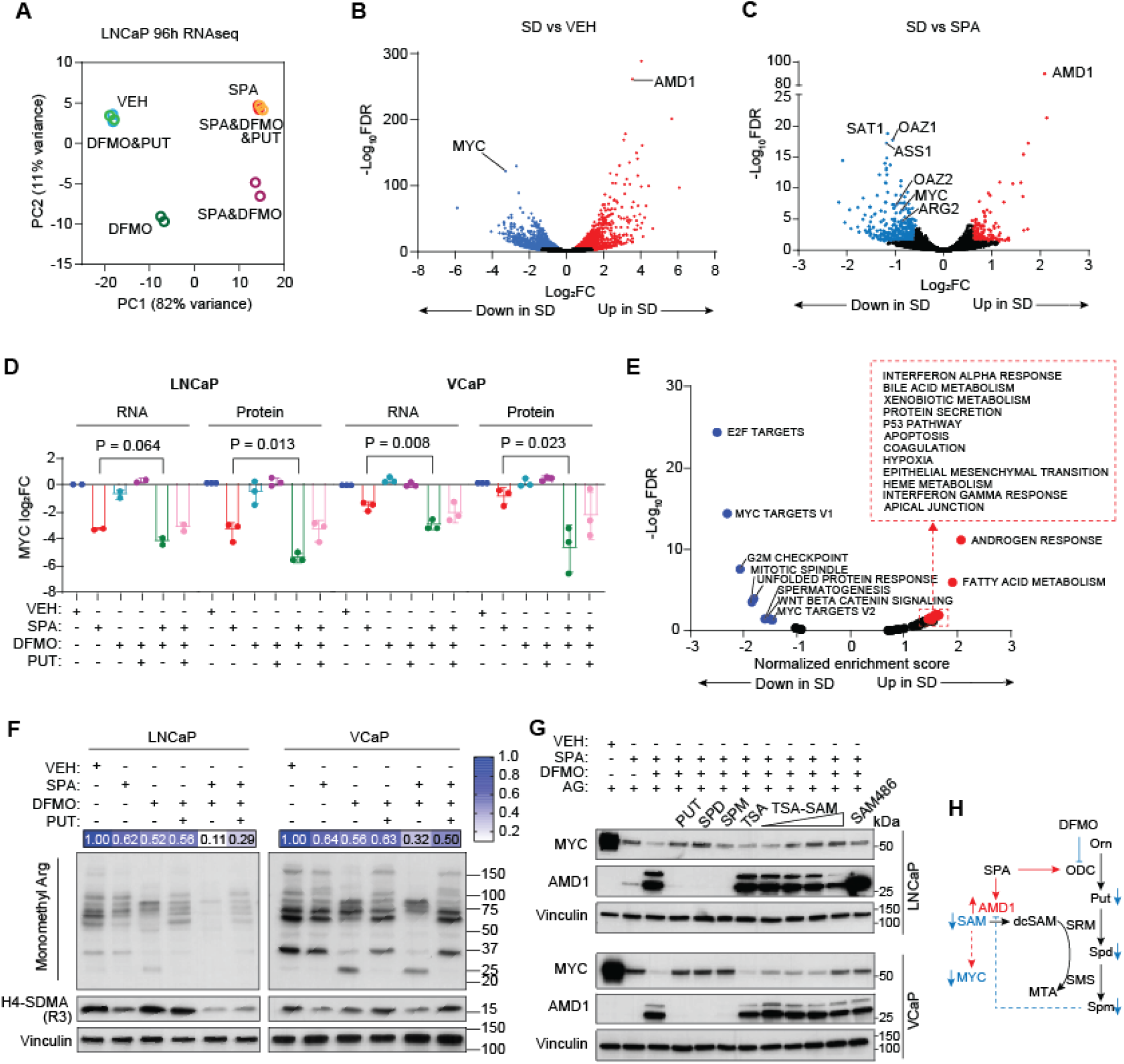
Inhibition of ODC increases downregulation of MYC by SPA by depleting S-adenosylmethionine. **A**. Principal component analysis of RNA transcript abundance by RNAseq of LNCaP cells treated with vehicle control (VEH; EtOH 0.1%), supraphysiological androgen (SPA, R1881 10nM), DFMO (5mM), DFMO and putrescine (PUT; 100uM), DFMO and SPA (SD), and DFMO and SPA and PUT for 96 hours. **B.** Volcano plot displaying change in transcript abundance in LNCaP cells treated with DFMO and SPA (SD) versus VEH as per (A). **C.** Volcano plot displaying change in transcript abundance in LNCaP cells treated with DFMO and SPA versus SPA as per (A). **D.** RNA and protein expression of MYC by RT-PCR and western blot of LNCaP and VCaP cells treated with vehicle control (VEH; EtOH 0.1%) or combinations of supraphysiological androgen (SPA, R1881 10nM), DFMO (5mM), and putrescine (PUT; 100uM), as indicated, for 96 hours. *MYC* RNA expression was normalized to *ACTB* and MYC protein expression was normalized to vinculin. P values by unpaired 2-tailed t test. **E.** Volcano plot displaying normalized enrichment scores of HALLMARK gene sets in LNCaP cells treated with DFMO and SPA versus VEH as per (A). **F.** Abundance of proteins with monomethyl arginine and H4-SDMA in LNCaP and VCaP cells following treatment with vehicle control (VEH; EtOH 0.1%) or combinations of supraphysiological androgen (SPA; R1881 10nM), DFMO (5mM), and putrescine (100uM) as indicated for 96 hours. Vinculin is used as a loading control. Representative blots of n = 2 independent experiments. **G**. Protein expression of MYC and AMD1 by western blot in LNCaP and VCaP cells treated with vehicle control (VEH, EtOH 0.1%) or combinations of supraphysiological androgen (SPA; R1881 10nM), DFMO (5mM), putrescine (PUT; 100uM), spermidine (SPD; 100uM), spermine (SPM; 100uM), toluenesulfonic acid (TSA; 200uM), S-adenosyl-methionine-toluenesulfonic acid (TSA-SAM; 25uM, 50uM, 100uM, 200uM), and SAM486 (2uM), as indicated, for 96 hours. All samples were treated with aminoguanidine (AG; 1mM). Vinculin is used as a loading control. **H.** Schematic displaying effects of SPA and DFMO combination treatment. SPA increases expression of ODC and AMD1. DFMO inhibits ODC activity, leading to depletion of downstream polyamines. Decreased polyamines reduces negative regulation of AMD1, further increasing its abundance. This marked increased in AMD1 by SPA and DFMO leads to depletion of SAM, which leads to decreased MYC expression.

Next, we assessed which specific transcripts are altered by SPA and DFMO combination therapy. Notably, *MYC* was markedly reduced by SPA and DFMO, to a greater extent than either monotherapy, which occurred in multiple cell lines, and translated to reduced MYC protein abundance and expression of MYC target genes (**Fig 6B-D**). This is notable because MYC is a key driver of PCa (*10, 36, 37*), and its downregulation contributes to efficacy of SPA monotherapy (*8*). MYC was also more suppressed by SPA in LNCaP cells expressing dCas9-KRAB blocking induction of *ODC1* compared with parental LNCaP cells, which was rescued by constitutive overexpression of ODC (**Fig S6B**), indicating that ODC induction serves to partially maintain MYC upon SPA treatment.

We also observed that SPA and DFMO markedly increased abundance of *AMD1* (**Fig 6B-C**). While SPA monotherapy also increased *AMD1,* as expected based on results shown in Fig 2B, *AMD1* abundance was much higher in cells treated with SPA and DFMO (**Fig S6D**). It was previously shown that polyamine depletion by DFMO increases abundance of *AMD1* transcript in prostate tissue (*38*), presumably as a feedback loop to regulate polyamine abundance. Actinomycin D chase experiments suggested that the 5-fold increase in *AMD1* transcript by DFMO was due at least in part to its increased mRNA stability, as the half-life of *AMD1* increased from about 1 hour in LNCaP cells treated with vehicle control to greater than 8 hours in cells treated with DFMO (**Fig S6E**).

We hypothesized that the increased abundance of *AMD1* by SPA and DFMO may lead to increased activity of AMD1, depletion of its substrate SAM, and alterations in epigenetic regulation of *MYC* transcription related to decreased bioavailability of methyl donors. To assess global protein methylation, we measured total monomethylated arginine and dimethylated lysine by western blot. In congruence with the decreased abundance of ADMA and SDMA in SKCaP tumors treated with SPA (**Fig 1B-C**), we observed that SPA monotherapy decreased abundance of total monomethylated arginine and dimethylated lysine in LNCaP and VCaP cells (**Fig 6F** and **Fig S6F).** DFMO monotherapy similarly decreased protein methylation, which could be reversed by putrescine (**Fig 6E** and **Fig S6F**). Interestingly, monomethylated arginine and H4-SDMA were most reduced by the combination of SPA and DFMO, more so than either monotherapy (**Fig 6E**). This confirms that each monotherapy and the combination therapy globally reduce protein methylation. Importantly, this decrease in methyl donors was found to be upstream of downregulation of MYC, as decreased MYC by SPA and DFMO could be rescued to the level of SPA monotherapy by multiple interventions, including (1) adding back polyamines, which reduced AMD1, (2) adding back SAM (provided as TSA-SAM with TSA salt used as a control compound), or (3) by inhibiting AMD1 with SAM486 (**Fig 6F**). This suggests that ODC inhibition enhances suppression of MYC by SPA by stimulating AMD1 leading to depletion of SAM (**Fig 6G**).

Notably, downregulation of MYC was not required for enhanced growth inhibition by SPA and DFMO, as constitutive expression of MYC in LNCaP cells did not rescue this growth defect (**Fig S6G-H)**. While a decrease in MYC function highly restricts growth of PCa (*39*), these data suggest that SPA and DFMO decreases PCa proliferation through multiple mechanisms. A possibility is that the global reduction of protein methylation by SPA and DFMO affects multiple signaling pathways and is not specific to MYC.

### DFMO decreases polyamines in patients with PCa treated with BAT

To assess whether DFMO enhances efficacy of BAT in patients with metastatic castration-resistant PCa, we designed a clinical trial, entitled the APEX trial (<u>A</u>ndrogen and <u>P</u>olyamine <u>E</u>limination alternating with <u>X</u>tandi; NCT06059118). Eligible patients are those with metastatic PCa that has progressed following treatment with an AR pathway inhibitor who lack significant pain due to their PCa. Patients are treated with a DFMO monotherapy lead-in phase followed by combination treatment with DFMO and BAT alternating with enzalutamide. The trial is actively enrolling, and we will report safety and efficacy endpoints when the prespecified number of patients are assessed.

For this trial, we selected a dose of DFMO of 1000 mg by mouth twice daily without adjustment for body surface area. This dose is comparable to the dose that was recently found to be safe and effective and approved for use by the US Food and Drug Administration as maintenance therapy after immunotherapy for children with high-risk, recurrent neuroblastoma (*40*). Further, Simoneau et al. previously found that DFMO 500 mg/m^2^ by mouth daily reduced polyamine abundance in the normal prostate over 28 days, with a median decline in putrescine of 95%, in spermidine of 69%, and in spermine of 55% (*41*). A follow-up larger study suggested that lower dose DFMO 500 mg by mouth daily (no body surface area adjustment) reduced putrescine abundance in the normal prostate over 12 months (median decline of 61%), but did not alter abundance of spermidine nor spermine (*42*). Given our data that ODC is induced by AR, which is more active in metastatic PCa than the normal prostate and is further activated when testosterone is administered, we opted for a higher dose of 1000 mg twice daily, more closely aligning with practice in the treatment of neuroblastoma.

To confirm that this dose of DFMO is sufficient to reduce polyamines despite testosterone treatment in patients with metastatic PCa, we measured plasma metabolites in the first 5 patients on trial, as well as in four age- and sex-matched healthy controls (HC). The patients had metastatic PCa that has progressed through multiple prior systemic therapies (range 4-8) with a complement of genomic features typical of advanced prostate cancer and a median prostate specific antigen (PSA) level of 255 ng/ml (range 10-2,387) (**Fig 7A**). Blood was collected pre-treatment on day 1, following DFMO monotherapy on day 8, and following DFMO and testosterone combination therapy on day 64 (**Fig 7B**). Across 414 measured metabolites, these samples clustered primarily by patient and disease state (i.e., HC versus patient) by principal component analysis (**Fig 7C**) and Euclidean clustering (**Fig S7A**). This indicates that patient-specific factors, rather than treatment, are the primary determinants of metabolite composition of these samples.

**Fig. 7.**
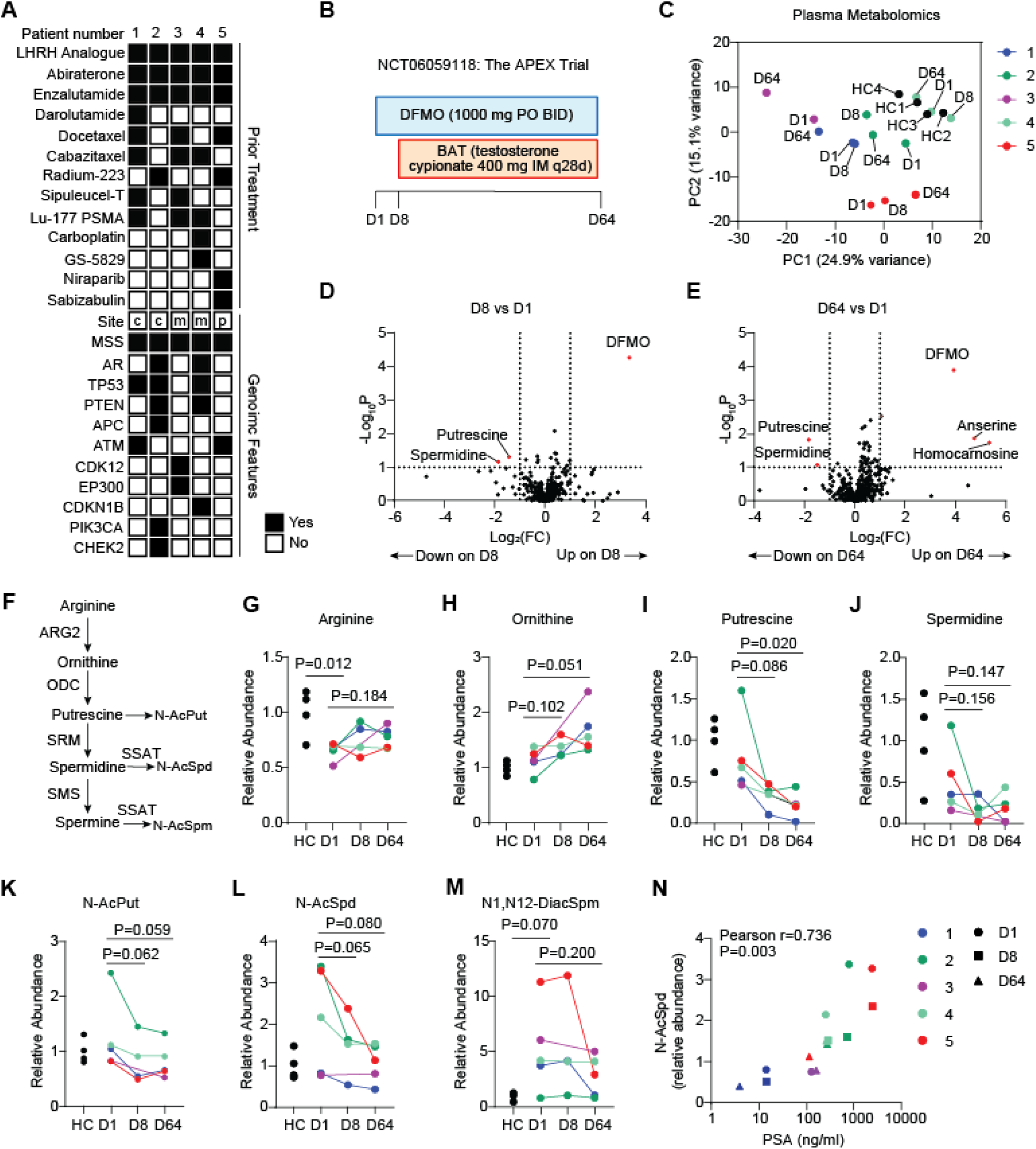
DFMO decreases polyamines in patients with PCa treated with BAT. **A.** Patient characteristics including prior treatments and tumor genomic features for first 5 patients on trial. For site, c indicates circulating tumor DNA, m indicates metastatic tumor site, and p indicates primary tumor. **B.** Trial schematic for days 1-64. Participants are treated with DFMO monotherapy from day 1 to day 8, then DFMO and testosterone from day 9 to day 64. D, day. **C.** Principal component analysis of fasting metabolite abundance in plasma of the first five patients on trial at indicated timepoints and 4 healthy controls (HC). **D.** Volcano plot displaying log_2_ fold-change (Log2(FC)) of metabolites in plasma on day 8 versus day 1. **E.** Volcano plot displaying log_2_ fold-change (Log2(FC)) of metabolites in plasma on day 64 versus day 1. **F.** Schematic of metabolites involved in de novo polyamine synthesis regulated by AR. **G-M**. Relative abundance of arginine (**G**), ornithine (**H)**, putrescine (**I**), spermidine (**J**), N-acetylputrescine (**K**), N-acetylspermidine (**L**), and N1,N12-diacetylspermine (**M**) in plasma of HC and trial participants at indicated timepoints. P values comparing HC and D1 by unpaired 2-tailed t test. P values comparing D1 and D8 or D1 and D64 by paired 2-tailed t test. **N.** Correlation of N-acetylspermidine and PSA. R and p values determined by Pearson’s correlation calculation.

Nonetheless, when comparing D8 plasma versus D1 plasma, putrescine and spermidine abundance were decreased in concert with an increase in DFMO (**Fig 7D**). Importantly, putrescine and spermidine abundance were persistently reduced on D64, despite testosterone treatment and its likely induction of ODC (**Fig 7E**). The administration of testosterone had no effect on abundance of plasma DFMO between D8 and D64 (**Fig S7B**). These unbiased analyses revealed that two analogs of carnosine, anserine and homocarnosine, were markedly increased in plasma on D64 (**Fig 7E** and **Fig S7C-D**). Anserine and homocarnosine are known to be highly abundant in cardiac and skeletal muscle. They possess functions including pH-buffering, metal-ion chelation, and antioxidant capacity, and have previously been shown to restrict growth of cancer models (*43*), but their regulation and function are unclear in our patients.

Next, we specifically assessed changes to metabolites involved in de novo polyamine synthesis (**Fig 7F**). Interestingly, arginine was lower in plasma of patients pre-treatment versus HCs and increased in several patients following treatment with DFMO (**Fig 7G**). Ornithine abundance was similar in plasma of patients pre-treatment versus HCs, but also increased following treatment with DFMO (**Fig 7H**). Putrescine, spermidine, N-acetylputrescine, and N-acetylspermidine abundance was similar in plasma of patients pre-treatment versus HC, but in contrast to arginine and ornithine, levels fell following treatment with DFMO on D8 and were persistently lowered on D64 (**Fig 7I-L**). Spermine was not detectable by our methods, however N1,N12-diacetylspermine was detected and notable for higher levels in patients pre-treatment compared with HC and significant decline in two patients on D64 (**Fig 7M**). The observed decrease in plasma polyamines might be due to decreased tumor burden due to therapy, leading to decreased release from tumor to the bloodstream, rather than decreased synthesis due to effective inhibition of ODC by DFMO. Serum PSA is a tumor marker that is used to approximate tumor burden in PCa. Notably, there was no correlation between putrescine, spermidine, or N-acetylputrescine with serum PSA (**Fig S8A-E**), suggesting that their decline was primarily driven by decreased rate of synthesis. Interestingly, there was a strong positive correlation between PSA and N-acetylspermidine across timepoints, including the pre-treatment D1 timepoint (**Fig 7N**). This suggests that plasma N-acetylspermidine may be primarily derived from tumor and, like PSA, act like as a tumor marker to reflect tumor burden and AR activity. Nonetheless, these data suggest that DFMO at a dose of 1000 mg by mouth twice daily can effectively inhibit ODC activity, resulting in accumulation of its precursors and depletion of its products, putrescine and spermidine, in patients with metastatic castration-resistant PCa treated with testosterone.

## DISCUSSION

The AR is widely appreciated to regulate cellular metabolism. Here we demonstrate that the AR alters metabolism of PCa through induction of de novo polyamine synthesis from arginine (**Fig S9A**). The polyamines, putrescine, spermidine, and spermine are small polycationic alkylamines present in millimolar concentrations in mammalian cells (*11*). Our results indicate that AR increases their synthesis through regulation of multiple enzymes involved in this pathway including ODC, AMD1, ARG2, and SMS. We delineate the mechanism by which AR regulates ODC to be through direct AR DNA binding at enhancer sites upstream of the *ODC1* promoter. Interestingly, we found that these sites were also bound by AR in normal prostate tissue, which has the highest abundance of spermine of any measured non-malignant tissue, suggesting that the mechanism by which AR increases polyamines is conserved between normal and malignant prostate. We further found that AR inhibition reduced ODC in activated human T lymphocytes, cells of distinct developmental origin than prostate epithelia. Altogether, the data suggest that AR is a fundamental regulator of ODC and polyamine synthesis in diverse mammalian cell types.

Polyamines have wide-ranging effects on cellular function by modifying intracellular as well as intercellular signaling that tends to enhance cell survival, proliferation, and resilience under stress (*24*). As such, these metabolites support several hallmarks of cancer, and polyamine synthesis is known to be activated in diverse cancer types (*11, 44*). In cancer, several common oncogenic signaling pathways were previously known to drive polyamine synthesis, including those driven by RAS (*45–47*), PI3K (*48*), and perhaps most notoriously, MYC (*31, 49*). Our data suggest that polyamine synthesis is driven primarily by the AR in prostate cancer and highlight a subtlety to MYC regulation of ODC. In cells with high AR, MYC did not induce ODC, but in fact antagonized AR-mediated induction of ODC to reduce ODC abundance. This may occur through competition for transcriptional co-factors and indicate that AR is a more active positive regulator of ODC, such that transcriptional interference by a less potent activator, MYC, reduces AR target gene expression in prostate cancer. As such, we anticipate that novel agents that target MYC will be unlikely to reduce polyamine synthesis in this context, and may actually enhance polyamine synthesis, in AR-positive PCa in the absence of co-targeting the AR.

The objective of this work was to identify metabolic vulnerabilities induced by SPA. We show that dCas9-KRAB-mediated inhibition of AR regulation of ODC or direct ODC inhibition by DFMO enhanced efficacy of SPA in PCa models. This implies that AR stimulation of polyamine synthesis facilitates fitness of PCa treated with SPA. Prior work has highlighted the varied roles of polyamines in driving cancer progression (*11, 44, 50*). Here, we demonstrate a less appreciated function of polyamines to negatively feed-back on AMD1 to temporize its activity and depletion of its substrate SAM (**Fig S8B**). SAM is the major methyl donor for protein methylation in mammalian cells, and its abundance is known to alter histone methylation and epigenetic regulation of gene expression (*51*). We found that SPA and DFMO led to global protein demethylation and enhanced suppression of the proto-oncogene *MYC*. MYC suppression appeared to be due to polyamine depletion leading to increased AMD1 activity and depletion of SAM because it could be rescued by addition of polyamines, AMD1 inhibition, or addition of SAM. A prior study established precedent that SAM depletion can decrease proliferation of PCa, in which SAM depletion was induced by stimulating polyamine catabolism concurrent with inhibition of the methionine salvage pathway (*52*). Our data suggest that AMD1 is an important regulator of SAM abundance in PCa and supports a growing appreciation that metabolism can regulate oncogenic cell signaling (*53*).

It is important to note that while MYC is a critical driver of advanced prostate cancer, our data indicate that its downregulation is not required for growth inhibition by SPA and DFMO. This suggests that multiple growth-promoting pathways are affected by the combination treatment, in addition to those regulated by MYC. Additionally, our work focused on cancer-cell intrinsic effects of treatment. Yet we found that SPA enhanced both intracellular abundance and extracellular secretion of polyamines by PCa. It is possible that these secreted polyamines have important effects on intratumoral stromal and immune cells in tumors of patients treated with BAT. Spermine and spermidine dampen production of pro-inflammatory cytokines in macrophages and drive differentiation and survival of immune-suppressive myeloid cells (*54–60*). Polyamines are also required for T cell lineage commitment and restraint of T cell inflammatory cytokine production, with deletion of *ODC1* in T cells driving robust production of IFNγ and autoimmune colitis in mice (*61*). In models of colon cancer, glioblastoma, breast cancer, and melanoma, inhibition of polyamine synthesis enhanced tumor immunity and augmented efficacy of anti-PD1 administration (*59, 62, 63*). Assessment of changes to tumor-infiltrating immune cell abundance and phenotype is a key secondary objective of the described ongoing clinical trial of BAT in combination with DFMO for patients with metastatic castration-resistant PCa (NCT06059118).

To ensure adequate dosing of DFMO in this trial, we performed pharmacodynamic studies for the first 5 patients on trial. We observed that DFMO resulted in accumulation of ornithine and reduction of putrescine and spermidine in plasma prior to and following treatment with BAT. This suggests that the dosing of DFMO is adequate to inhibit ODC in these patients, even despite ODC induction by AR, and the trial is adequately designed to assess its primary objective to determine whether inhibition of ODC by DFMO enhances efficacy of BAT in patients with metastatic PCa.

Another notable finding of the plasma metabolomic studies was the reduction of arginine in plasma of patients with PCa relative to HCs. Tumor arginine was previously demonstrated to be even lower than plasma arginine in murine cancer models (*64*), which has been attributed to high activity of ARG1 of tumor-infiltrating suppressive myeloid cells (*65*). We observed a trend toward increased plasma arginine with DFMO treatment, suggesting that arginine depletion may also be due in part to its utilization for polyamine synthesis. This is notable because arginine depletion has been described to decrease T cell survival and antitumor cytotoxic function (*66*), implying that increasing arginine in the tumor microenvironment may enhance tumor immunity. In these data, we also identified a positive correlation between serum PSA and acetylated spermidine, which was present in pre-treatment samples and persistent in on-treatment samples. This suggests that acetylated spermidine in plasma may be primarily derived from tumor, with its abundance primarily determined by tumor volume. We suspect that acetylated spermidine is a redundant biomarker with PSA without a distinct advantage as a tumor marker.

There are several limitations to this study. First, we have primarily studied AR regulation of polyamine synthesis in contexts in which SPA monotherapy is growth-suppressive. In these contexts, ODC activation appears to be a mechanism to resist the growth-suppressive effects of SPA, exemplified by its positive regulation of MYC expression. The functional role of stimulated ODC in contexts in which SPA monotherapy fails to suppress growth is unclear. While cells with primary resistance to SPA (LAPC4 and 22Rv1) were growth-suppressed by DFMO, we did not observe enhanced effect by combination treatment with DFMO and SPA, suggesting that enhanced polyamine synthesis is not the primary mechanism of resistance to SPA in these cell lines. We previously demonstrated that the primary mechanism of resistance to SPA in these cell lines is low AR activity (*8*). Secondly, we have focused on cancer-cell intrinsic effects of treatment despite evidence that SPA increases polyamine secretion and very well may alter function of surrounding cells in tumor. We hope to begin to address this question through study of paired metastatic biopsies of patients enrolled on our trial. Lastly, we have not delineated the specific mechanism by which depletion of SAM leads to downregulation of MYC. Future studies will delineate how bioavailability of SAM alters MYC expression.

DFMO is an old drug that has been extensively tested clinically, including some studies for PCa. It was one of the first non-heavy metal drugs to be approved for treatment of trypanosomiasis (i.e., African sleeping sickness) and is known to be extremely well-tolerated, even at high doses (*67, 68*). The first phase I trial of DFMO was performed at Johns Hopkins in 1984 for patients with advanced solid tumors or lymphomas (*69*), and shortly thereafter another phase I trial of DFMO and methylglyoxyl bis-(guanylhydrazone) (MGBG; an inhibitor of AMD1) was performed in 1986 and included 5 patients with PCa (*70*). The combination of DFMO and MGBG was toxic and ineffective for these patients. Notably our data suggests that AMD1 inhibition by MGBG may have counteracted tumor-suppressive effects of DFMO in this trial. The lack of efficacy observed in this trial stifled enthusiasm for further clinical development of DFMO as a treatment for advanced PCa. Nearly 40 years later, we believe our data support renewed enthusiasm to test efficacy of DFMO as treatment for patients with advanced PCa in a new context of combination therapy with BAT.

## MATERIALS AND METHODS

### Patient-derived xenograft (PDX) mouse studies

Male NSG mice aged 8-12 weeks were obtained from the Sidney Kimmel Comprehensive Cancer Center (SKCCC) Animal Core Facility and surgically castrated. SKCaP patient-derived xenograft tissue was minced, mixed with Matrigel (BD Biosciences) and implanted subcutaneously on the flank. Testosterone was administered by implantation of a slow-release subcutaneous pellet in the opposite flank. Pellets were assembled using 25-mm sections of Silastic Laboratory tubing, filled with 30 mg testosterone cypionate (Steraloids Inc A6960-000), sealed on both ends with Silastic Medical Adhesive Type A, then sterilized. Tumors were measured twice weekly using microcalipers, and tumor volume was calculated using the following formula: 0.5236 x L x W x H. At study completion, mice were euthanized and tumors were extracted and flash frozen and stored at −80°C. All mice were housed in the Johns Hopkins animal facility and we have complied with all relevant ethical regulations in accordance with Johns Hopkins Institutional Animal Care and Use Committee.

### PDX global metabolomics

Frozen tumor specimens were shipped to Human Metabolome Technologies, Inc. Samples were mixed with 50% acetonitrile in water containing internal standards with volume normalized to tissue weight and homogenized (1,500 rpm, 120 sec x 5 times). 400ul of supernatant was filtered through a 5-kDa filter (ULTRAFREE-MC-PLHCC, Human Metabolome Technologies), then centrifugally concentrated and resuspended in 50ul ultrapure water immediately before measurement. Samples were measured in the cation and anion modes of Capillary Electrophoresis Time-of-Flight Mass Spectrometry (CE-TOFMS) with 1:3 dilution for anion mode. Cation Mode conditions were as follows. Device: Agilent CE-TOFMS system (Agilent Technologies Inc) Machine No. 6; Capillary, fused silica capillary i.d. 50um x 80cm. Analytical Condition: Run buffer, Cation Buffer Solution (p/n: H3301-1001); Rinse Buffer Solution (p/n: H3301-1001); Sample Injection, Pressure injection 50mbar, 10 sec; CE voltage, Positive 30kV; MS ionization, ESI positive; MS capillary voltage, 4,000V; MS scan range, m/z 50-1,000; Sheath liquid, HMT Sheath Liquid (p/n: I3301-1020). Anion Mode conditions were as follows. Device: Agilent CE-TOFMS system (Agilent Technologies Inc) Machine No. 2; Capillary, fused silica capillary i.d. 50um x 80cm. Analytical Condition: Run buffer, Anion Buffer Solution (p/n: H3302-1023); Rinse Buffer Solution (p/n: H3302-1023); Sample Injection, Pressure injection 50mbar, 22 sec; CE voltage, Positive 30kV; MS ionization, ESI negative; MS capillary voltage, 3,500V; MS scan range, m/z 50-1,000; Sheath liquid, HMT Sheath Liquid (p/n: I3301-1020). Peaks detected in the CE-TOFMS analysis were extracted using automatic integration software (MasterHands version 2.17.1.11 developed at Keio University) to obtain peak information, including m/z, migration time, and peak area. The peak area was then converted to relative peak area by dividing peak area by (internal standard peak area x sample amount). The peak detection limit was determined based on signal-noise ratio; S/N=3. Putative metabolites were then assigned from HMT’s standard library on the basis of m/x and MT. The tolerance was +/-0.5 min in MT and +/-10 ppm in m/z.

### Plasma global metabolomics

Whole blood was collected in EDTA vacutainer tubes then transferred to 15-ml conical tube and centrifuged at 2400 RPM for 5 minutes at room temperature. The top plasma layer was transferred to a new 15-ml conical tube and centrifuged at 8000 RPM for 5 minutes at room temperature. The top plasma layer was then aliquoted into cryovials and stored at −80°C. Plasma samples were shipped to Human Metabolome Technologies, Inc. Each 50-ul sample was mixed with 200ul methanol containing internal standards and 150ul of Milli-Q water, then filtered at 4°C through a 5-kDa filter (ULTRAFREE-MC-PLHCC, Human Metabolome Technologies, Yamagata, Japan). The filtrate was centrifugally concentrated and resuspended in ultrapure water for metabolome analysis. Samples were measured in the cation and anion modes of Capillary Electrophoresis Fourier Transform Mass Spectrometry (CE-FTMS) with 1:3 dilution for anion mode. Cation Mode conditions were as follows. Device: CE, Agilent CE system; MS, Q Exactive Plus Machine No.1; Capillary, fused silica capillary i.d. 50um x 80cm. Analytical Condition: Run buffer, Cation Buffer Solution (p/n: H3301-1001); Rinse Buffer Solution (p/n: H3301-1001); Sample Injection, Pressure injection 50mbar, 10 sec; CE voltage, Positive 30kV; MS ionization, ESI positive; MS capillary voltage, 4,000V; MS scan range, m/z 60-900; Sheath liquid, HMT Sheath Liquid (p/n: I3301-1040). Anion Mode conditions were as follows. Device: CE, Agilent CE system; MS, Q Exactive Plus Machine No.5; Capillary, fused silica capillary i.d. 50um x 80cm. Analytical Condition: Run buffer, Anion Buffer Solution (p/n: H3302-1023); Rinse Buffer Solution (p/n: H3302-1023); Sample Injection, Pressure injection 50mbar, 25 sec; CE voltage, Positive 30kV; MS ionization, ESI negative; MS capillary voltage, 3,500V; MS scan range, m/z 70-1,050; Sheath liquid, HMT Sheath Liquid (p/n: I3301-1040). Peaks detected in the CE-FTMS analysis were extracted using automatic integration software (MasterHands version 2.19.0.2 developed at Keio University) to obtain peak information, including m/z, migration time, and peak area. The peak area was then converted to relative peak area by dividing peak area by (internal standard peak area x sample amount). The peak detection limit was determined based on signal-noise ratio; S/N=3. Putative metabolites were then assigned from HMT’s standard library on the basis of m/x and MT. The tolerance was +/-0.5 min in MT and +/-5 ppm in m/z.

### Metabolomics Analysis

Metabolomics analysis was performed in MetaboAnalyst version 6.0 (*71*). Data was filtered by removing 20% with smallest interquartile range, log transformed, auto-scaled, and visualized by principal component analysis (PCA), hierarchical clustering dendrogram with Euclidean distance and Ward clustering algorithms. Differentially abundant metabolites with greater than 2-fold change and lower than 0.1 p value was assessed between conditions.

### Cell Culture

LNCaP and VCaP cell lines were obtained from American Type Culture Collection (ATCC). LAPC4 and 22Rv1 cell lines were a gift from J Isaacs. LNCaP, LAPC4, and 22Rv1 were grown in RPMI 1640 (Gibco; 11835-055) supplemented with 10% fetal bovine serum (Corning), sodium lactate 1.6mM, sodium pyruvate 0.5mM, L-alanine 0.43mM, 1% pen-strep (Gibco). VCaP was grown in DMEM (ATCC 30-2002) supplemented with 10% fetal bovine serum (Corning) and 1% pen-strep (Gibco). The vectors pCDH-puro-cMyc (Addgene; 46970, J Wang laboratory), pCDH-EF1-FHC empty vector control (Addgene, 64874, R Wood laboratory), piSMARTvector-PGK-TurboGFP-TRE3G-shAR vectors (Horizon; V3IHSPGG_8216343 and V3IHSPGG_7292640), pLX303-ZIM3-KRAB-dCas9 (Addgene; 154472), or pLV[Exp]-Neo-CMV-hODC1 vector (VectorBuilder; VB240830) were transfected into 293T cells (ATCC) along with pMD2.G (Addgene; 12259) and psPAX2 (Addgene; 12260) packaging vectors using lipofectamine 2000 (Invitrogen) to produce cMyc-puro, control-puro, Tet-On-shAR-puro, KRAB-dCas9-blasti, and ODC-Neo lentivirus particles. Two days after transduction with indicated virus, vector-expressing cells were selected with puromycin 1ug/ml for 72 hours, blasticidin 10ug/ml for 7 days, or neomycin 500ug/ml for 7 days. To produce cells with KRAB-dCas9 targeting ARBS upstream of *ODC1*, LNCaP-KRAB-dCas9 cells were transduced with sgRNAs targeting site 1:

A: GCTCCTCCAAAAGGATCTGCTGG
B: GCCAAATAGGAACTAGGCCAGG

or sgRNAs targeting site 2:

C: CCGGAACAAGTTGGCCCGCCAGC
D: GTGCGGAATATGTTTATGAAAGG

or a non-targeting sgRNA:

NT: CAACGTCGCGAACGTCGTAT

cloned into lenti-Guide-puro (Addgene; 52963) and selected with puromycin 1ug/ml for 72 hours. Cells were maintained at 37 ⁰C in 5% CO2. They regularly tested negative for mycoplasma contamination using MycSensor PCR Assay kit (Agilent Technologies). R1881, SAM486, putrescine, spermidine, spermine, TSA, TSA-SAM, and aminoguanidine were obtained from Sigma. DFMO from Professor Patrick Woster.

### Isotope Tracing

LNCaP or VCaP cells were plated in standard culture media at a density of 4 million cells in a 10-cm dish. The following day, media was exchanged for media containing either uniformly-labeled ^13^C-arginine (SILAC (Thermo Scientific 88365) with 1.1mM U-13C-arginine (Cambridge Isotope Laboratories, CLM-2265-H-PK) and 100uM unlabeled putrescine) or ^13^C-putrescine (RPMI, which contains 1.1mM unlabeled arginine, with 100uM U-13C-putrescine (Cambridge Isotope Laboratories, CLM-6574-PK)) with either vehicle control (EtOH 0.1%) or SPA (R1881 10nM). Following 24 hours, intracellular and extracellular metabolites were collected. For extracellular metabolites, media was collected, run through a 3kDa filter (Sigma UFC5003), and stored at - 80°C. For intracellular metabolites, cells were washed with PBS, then metabolites extracted in 80% methanol in water, samples flash-frozen in liquid nitrogen, and stored at −80°C. Supernatant was collected after centrifugation at 15,000g for 10 minutes and dried by a SpeedVac. Dried metabolite extracts were resuspended in 50% acetonitrile solution. LC-MS based metabolomics profiling was performed on an LC-MS system consisting of an Agilent 1290 Infinity II Binary UHPLC pump and a Bruker timsTOF Pro II mass spectrometer. HILIC-LC chromatographic separations were performed on the above UHPLC using a Waters XBridge BEH Amide column (2.1 x 150 mm, 1.7μm). The LC parameters were as follows: autosampler temperature, 4 °C; injection volume, 2 ml; column temperature, 40 °C; and flow rate, 0.20 ml/min. The solvents and optimized gradient conditions for LC were: Solvent A, Water with 0.1% formic acid; Solvent B, Acetonitrile with 0.1% formic acid; A non-linear gradient from 99% B to 45% B in 25 minutes with 5min of post-run time. MS spectra were collected using a timsTOF Pro II mass spectrometer (Bruker Daltonics) equipped with IonBooster ESI source. The mass spectrometer was operated in negative mode with auto MS/MS method. The optimized operation parameters were: End Plate Offset, 400V; Capillary Voltage,1000 V; Charging Voltage, 300 V; Nebulizer Pressure, 4.1 bar; Dry Gas 3 L/min; Dry Gas Temperature, 200 °C; Vaporizer Temperature, 350 °C; The mass scan range, 70-1100 m/z; Scan rate, 12 Hz. Spectra were internally mass calibrated at the beginning of every sample run by infusion of a small fragment of reference mass solution using a syringe pump connected to the sprayer feeding into the ESI source. Data were acquired with Compass HyStar 5.1 acquisition software and processed with TASQ 2022. The analyte database used for metabolite identification was developed in house with retention times based on the HILIC method.

### Western blot analyses

Cells or tissues were lysed with denaturing lysis buffer (Cell Signaling technology) containing protease and phosphatase inhibitors (Roche). Protein concentration was determined using the Pierce BCA Protein Assay kit (Thermo Scientific), and 5-20 ug of lysate was resolved on a SDS-PAGE gel and transferred to a nitrocellulose membrane. Membranes were blocked with 5% milk for 1 hour, then incubated with primary antibody overnight. Primary antibodies used were: anti-cMYC (Abcam, clone Y69, 1:1000 dilution), anti-AR (Santa Cruz, sc7305, 1:1000 dilution), anti-PSA (Dako, A0562, 1:1000 dilution), anti-ODC (CST; 52238, 1:000 dilution), anti-AMD1 (Proteintech 11052-1AP, 1:2000 dilution), anti-ARG2 (abcam; AB137069, 1:1000 dilution), anti-Hypusine (Millipore; ABS1064, 1:2000 dilution), anti-EIF5A (BD Biosciences; 26/eIF-5a, 1:000 dilution), anti-SDMA-H4(Arg3) (CST; 92326S, 1:000 dilution), anti-MMArginine (CST; 8015, 1:000 dilution), anti-DmeLysine (CST; 14117, 1:000 dilution), anti-vinculin (Millipore, clone V284, 1:2000 dilution). Anti-rabbit IgG, HRP-linked Antibody (CST, 7074S, 1:5000 dilution) and Anti-mouse IgG, HRP-linked Antibody (CST, 7076S, 1:5000 dilution) were used as secondary antibodies.

### Quantitation of polyamines and polyamine metabolic enzymes

Enzymatic activity assays for ODC and AMD1 were performed on quick-frozen tissue lysates by measuring the production of ^14^C-labelled CO_2_, as previously described (*72, 73*). HPLC-based quantitation of polyamines and their acetylated derivatives was performed following pre-column dansylation of samples and standards. 1,7-Diaminoheptane was used as an internal standard. Polyamines and diaminoheptane used in standards were purchased from Sigma. Derivatization and chromatography parameters were based on the method of Kabra (*74*). Polyamine and enzyme levels are presented relative to total cellular protein in the lysate, as determined using the BioRad Protein Assay.

### Human T cell studies

First, PBMCs were isolated from healthy donors, and stored at −80°C. PBMC isolation entailed a 400 x g centrifugation for 5 min to separate cells from plasma. Cells were resuspended in RPMI 1640, underlaid with Ficoll and centrifuged at 400 x g for 30 min without breaks at room temperature. The PBMC layer was collected, washed twice with RPMI 1640, and cryopreserved in fetal bovine serum + 10% DMSO. Subsequently, PBMCs were thawed at 37°C and immediately transferred to RPMI 1640 (Gibco; 11875-135) supplemented with 10% fetal bovine serum (Fisher Scientific; 26140079), glutamine 2 mM (Quality Biological; 118-084-721), and 1% pen-strept (Gibco). PBMCs were resuspended in new RPMI media, and incubated for 24 hours at 37°C. Next, T lymphocytes were magnetically isolated by negative selection (Miltenyi Biotec; 130-096-535). Isolated T lymphocytes were plated in the presence or absence of Human T-Activator CD3/CD28 Dynabeads (Gibco; 11132D) as per manufacturer’s instructions, and in the presence of enzalutamide 10 uM or vehicle control (EtOH 0.1%). After 24 hours of incubation, cell suspensions were collected, cells were washed with ice cold PBS, and cell pellets were flash frozen in liquid nitrogen for subsequent lysis for western blot.

### Proliferation, cell viability, and cell cycle analyses of cell lines

For proliferation and cell viability assays, cells were plated in triplicate on 6-well (0.1 x 10^6^ cells/well) or 12-well (0.05 x 10^6^ cells/well) plates and incubated with treatment for indicated duration. Cells were counted using a hemocytometer with viability assessed by trypan blue exclusion. For cell cycle analysis, cell pellets were resuspended in cold 70% ethanol at least overnight, then subsequently washed with PBS, and resuspended in 50 ug/ml propidium iodide (Sigma) and 100 ug/ml RNAse (Sigma) and run on the BD FACSCelesta flow cytometer with analysis using FlowJo software 10.4.2.

### Quantitative real-time polymerase chain reactions

RNA was extracted using the RNeasy kit (Qiagen) and cDNA generated using the high-capacity cDNA reverse transcription kit (ThermoFisher). RT-PCR were performed in triplicate using 500ug cDNA, 10ul TaqMan Gene Expression Master Mix (ThermoFisher), and 1ul 20x TaqMan Gene Expression Assay probe mix for *MYC* (Hs00153408_m1), *AMD1* (Hs00750876_s1), and *ACTB* (Hs01060665_g1) (ThermoFisher) on an ABI7500 RT-PCR System (ThermoFisher). Relative gene expression was determined by delta-delta CT.

### RNA sequencing

For RNA sequencing of cell lines, purified RNA was sent to Admera Health who performed library preparation with polyA selection, sequencing on the Illumina HiSeq 2500 with read length configuration of 2×150bp with 60 million reads per sample, and read mapping to GRCM38. Sequenced libraries were processed with deepTools (*75*), using STAR (*76*), for trimming and mapping, and featureCounts (*77*) to quantify mapped reads. Raw mapped reads were processed in R (Lucent Technologies) with DESeq2 (*78*) to generate normalized read counts to determine differentially expressed genes with greater than 2-fold change and lower than 0.01 adjusted p value. Gene set enrichment analysis was performed using fGSEA (*79*).

### AR Chromatin Immunoprecipitation (ChIP)

Cells were grown to the 5×10^6^ density in 100 mm plates and were treated with either vehicle or 10nM R1881 for 24h, washed with PBS, and fixed in adherent condition using 1% paraformaldehyde in 5mL PBS at room temperature for 10 minutes with gentle rocking. Fixation was quenched by adding 1mL of ice-cold glycine, followed by incubation at room temperature for 10 min. Cells were washed twice with ice-cold phosphate-buffered saline (PBS), pelleted at 2000 rpm for 5 minutes at 4°C, and lysed in SDS lysis buffer (50 mM Tris-HCl pH 8.1, 50 mM EDTA, 1 % SDS, with protease and phosphatase inhibitors added) to release chromatin. Samples were snap-frozen and stored at −80°C until use. Frozen chromatin was thawed on ice and sonicated using a Covaris sonicator. For 1 mL samples, settings were Power: 140, Intensity: 3, Bursts per second: 200, and Duration: 1500 seconds. Sheared chromatin was clarified by centrifugation at maximum speed for 10 minutes at 4°C, and the supernatant was transferred to fresh tubes. Chromatin equivalent to 1 million cells was diluted in ChIP dilution buffer (16.7 mM Tris-HCl pH 8.1, 167 mM NaCl, 1.2 mM EDTA, 1.1% Triton X-100, 0.01% SDS, protease inhibitors). Antibodies specific to the AR (CST# 5153) (10 μL per 1 mL of diluted chromatin) or equivalent Rabbit IgG control (CST# 2729S) were added, and samples were incubated overnight at 4°C with rotation. The following day, Protein A/G Dyna beads (Thermo) (50 μL per sample) were washed in ChIP dilution buffer (16.7 mM Tris-HCl pH 8.1, 167 mM NaCl, 1.2 mM EDTA, 1.1% Triton X-100, 0.01% SDS) and added to the samples for 2 hours at 4°C with rotation. Beads were then subjected to sequential washes with low salt wash buffer [0.1%SDS,1%TritonX-100, 2(mM) EDTA, 20(mM) Tris-HCl, pH8.1,150 (mM) NaCl], high salt wash buffer [0.1% SDS, 1%Triton X-100, 2(mM) EDTA, 20(mM) Tris-HCl, pH 8.1, 500(mM) NaCl], LiCl wash buffer [0.25 (M) LiCl, 1% NP40, 1% deoxycholate, 1(mM) EDTA,10 (mM) Tris-HCl, pH8.1] and 1x TE buffer [10(mM) Tris-HCl,1(mM) EDTA pH8.0]. Finally, the complexes were eluted in elution buffer (1% SDS, 0.1 M NaHCO3) at room temperature. Eluted chromatin was supplemented with 5 M NaCl and incubated at 65°C overnight to reverse crosslinking. DNA was purified using AMPure beads according to the manufacturer’s protocol (Beckman Coulter). Beads were washed twice with 80% ethanol, air-dried, and eluted in 0.1× TE buffer. DNA was quantified and used for qPCR using SYBR Green using primers spanning site 1 and site 2:

Site 1-Forward: CTCTGAGCAATGGTGGCATC
Site 1-Reverse: TTCTCTGCAACTGTGTGTGC
Site 2-Forward: GGTGAACCGGAACAAGTTGG
Site 2-Reverse: TGCTGAGGTTATGATGGTGTT

Percent of input was calculated to estimate the relative AR binding at each site.

### Actinomycin D chase

Cells were treated with vehicle or DFMO for 96 hours then actinomycin D (5 µg/mL) was added to inhibit transcription. RNA was extracted at 0, 1, and 8 hours post-actinomycin D treatment and the relative expression of *AMD1* mRNA was quantified by RT-qPCR as above.

### Clinical trial design and procedures

The APEX (Androgen and Polyamine Elimination alternating with Xtandi; NCT06059118) clinical trial is a single-arm, open-label phase II study of bipolar androgen therapy (BAT) in combination with eflornithine alternating with enzalutamide for patients with metastatic CRPC that had progressed on at least one novel androgen receptor-target therapy. This study was approved by the Institutional Review Board at Johns Hopkins, and all accrued patients provided written informed consent. Patients are treated with repeat cycles of eflornithine 1000 mg by mouth twice per day and BAT (testosterone cypionate 400 mg intramuscular every 28 days) alternating with enzalutamide 160 mg by mouth daily. Cycles of treatment are repeated until progression or unacceptable toxicity. Response is assessed by PSA every 28 days, and periodic CT chest, abdomen, and pelvis and technetium-99 bone scan.

## List of Supplementary Materials

Fig S1 to S9.

Full uncut western blots.

Excel File 1. SKCaP-PDX metabolomics data

Excel File 2. Plasma metabolomics data

Excel File 3. Spermine concentration across human tissues.

## Data Availability

All data produced in the present study are available upon reasonable request to the authors.

## Acknowledgments

We thank the patients and caregivers who are participating in the clinical trial described. We also thank the clinical staff who are assisting in running the clinical trial described.

## Funding

National Institutes of Health grant K08CA273167 (LAS)

Department of Defense grant HT94252310107 (LAS)

The Prostate Cancer Foundation Young Investigator Awards (RK, DES, and LAS)

Phi Beta Psi Charity grant IPN22120375 (LAS)

The Samuel Waxman Cancer Research Foundation (TMS and RAC)

Panbela Therapeutics (LAS)

## Author contributions

Conceptualization: SRD, LAS

Methodology: RK, DES, LZ, EAT, TMS, RAC, LAS

Investigation: RK, SJ, DES, VV, AK, SK, SLD, LZ, JF, CEH, AN, SK, EP, PA, SW, LAS

Analysis: RK, SJ, DES, PA, CD, ET, EAT, JTI, AMD, TMS, RAC, LAS

Visualization: RK, DES, LAS

Funding acquisition: LAS

Supervision: ET, EAT, JTI, ELP, TMS, RAC, LAS

Writing – original draft: LAS

Writing – review and editing: All authors

## Data and materials availability

RNA sequencing data have been deposited to NCBI’s Gene Expression Omnibus and are accessible through GEO Series accession number TBD. Data from metabolomics and spermine concentration across human tissues are available in the supplementary materials.

