## Supplement for "Androgen receptor drives polyamine synthesis creating a vulnerability for prostate cancer"

### Supplemental Figures

Figure S1. SPA increases intracellular and extracellular polyamines in prostate cancer cell lines.
Figure S2. SPA increases activity of ODC and AMD1 in prostate cancer cell lines.
Figure S3. SPA does not alter abundance of hypusinated eIF5A.
Figure S4. Characteristics of subjects in the Hamalainen autopsy study.
Figure S5. dCas9-KRAB blocks AR binding upstream of *ODC1*
Figure S6. Inhibition of ODC increases downregulation of MYC by SPA by depleting S-adenosylmethionine.
Figure S7. Plasma metabolite analyses of trial participants.
Figure S8. Correlation of metabolites and PSA.
Figure S9. Schematic summary of findings.

**Supplemental Figures**

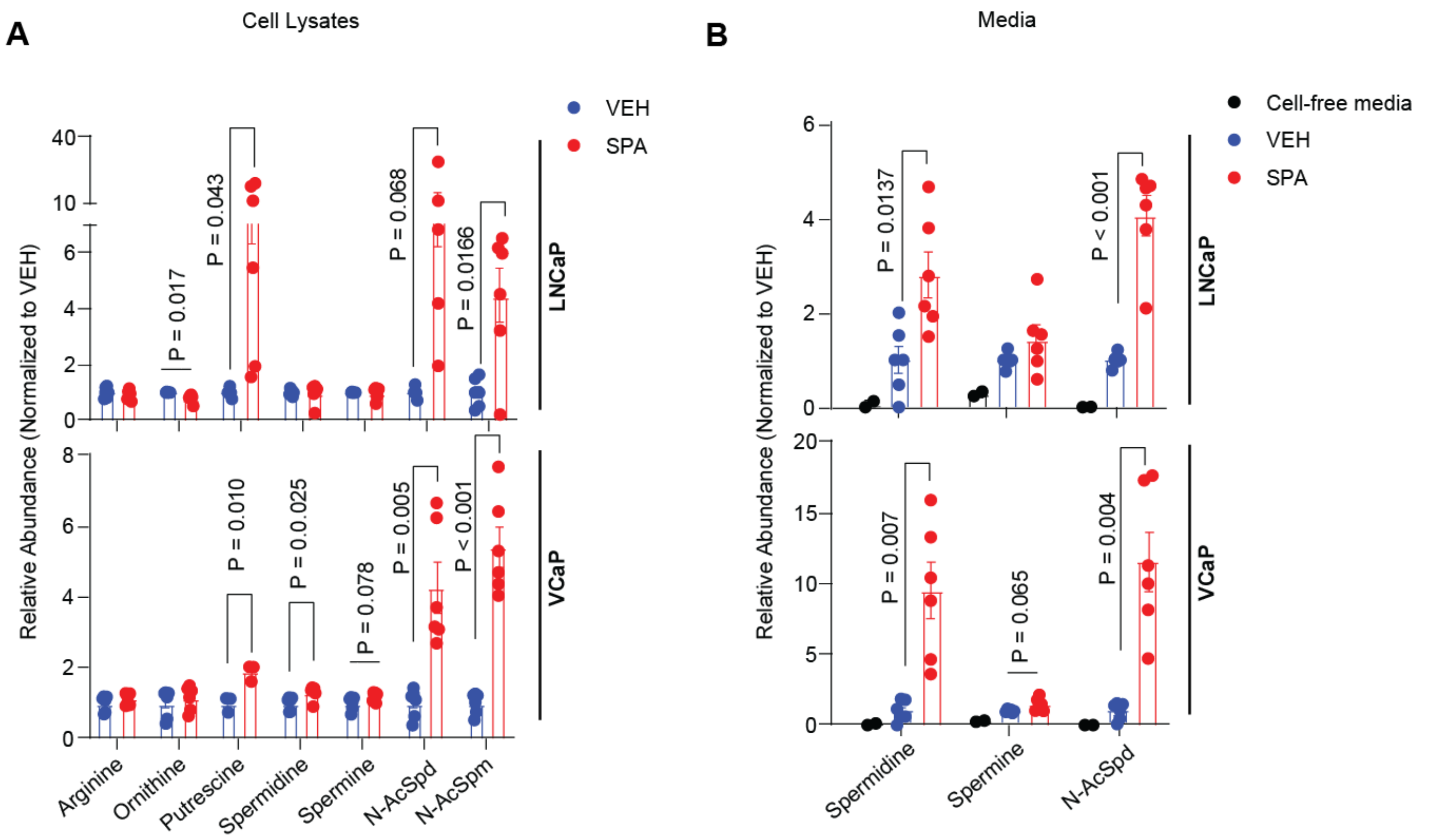


**Figure S1. SPA increases intracellular and extracellular polyamines in prostate cancer cell lines. A.** Relative abundance of indicated metabolites measured by LC-MS in LNCaP and VCaP cell lysates following 48 hours of treatment with vehicle control (VEH; EtOH 0.1%) or supraphysiological androgen (SPA; R1881 10nM). P values by unpaired 2-tailed t test. **B.** Relative abundance of indicated metabolites measured by LC-MS in LNCaP and VCaP media following 48 hours of treatment with VEH or SPA as per (A). P values by unpaired 2-tailed t test.

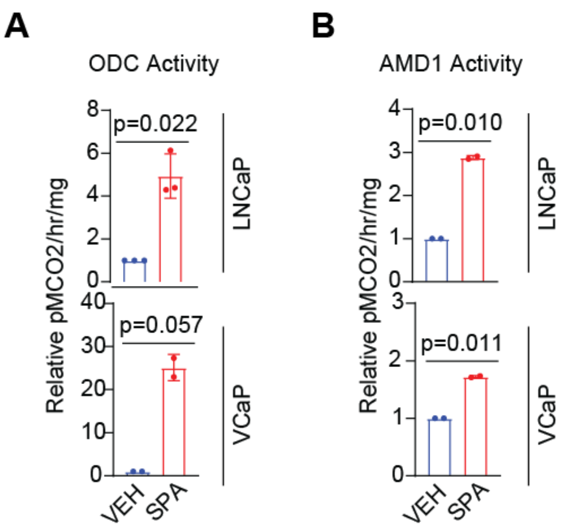


**Figure S2. SPA increases activity of ODC and AMD1 in prostate cancer cell lines. A.** ODC activity in cell lysates of LNCaP and VCaP cells treated with vehicle control (VEH; EtOH 0.1%) or supraphysiological androgen (SPA; R1881 10nM) for 24 hours. P values by unpaired 2-tailed t test. **B.** AMD1 activity in cell lysates of LNCaP and VCaP cells treated with vehicle control (VEH; EtOH 0.1%) or supraphysiological androgen (SPA; R1881 10nM) for 24 hours. P values by unpaired 2-tailed t test.

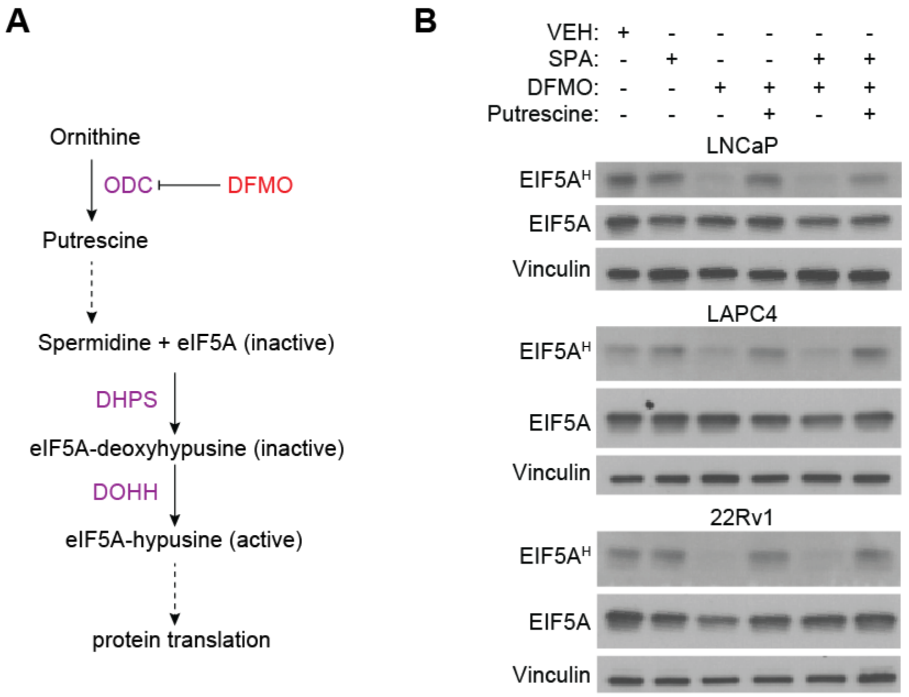


**Figure S3. SPA does not alter abundance of hypusinated eIF5A. A.** Schematic highlighting generation of hypusinated eIF5A from spermidine. **B.** Protein expression of hypusinated eIF5A ($EIF5A^H$) and total eIF5A in LNCaP, LAPC4, and 22Rv1 cells following treatment with vehicle control (VEH; EtOH 0.1%) or combinations of supraphysiological androgen (SPA; R1881 10nM), DFMO (5mM), and putrescine (100uM) as indicated for 96 hours. Vinculin is used as a loading control. Representative blots of n = 2 independent experiments.

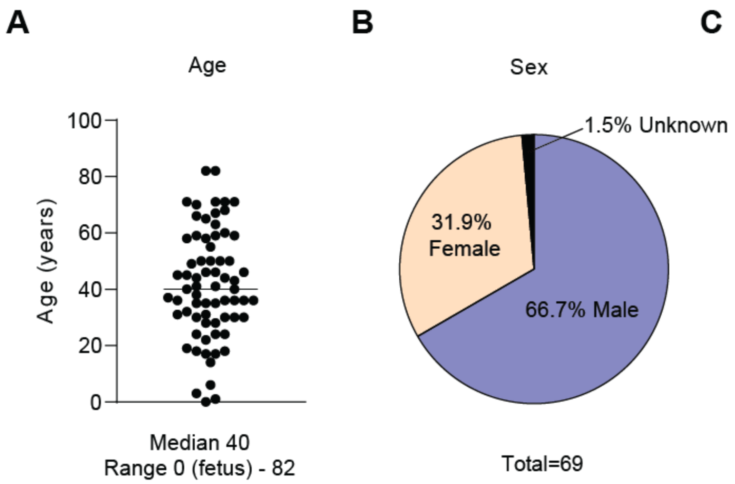


C

| Cause of Death | Number (Percent) |
|---|---|
| Trauma | 26 (37.7) |
| Cardiovascular disease | 9 (13) |
| Myocarditis | 4 (5.8) |
| Diabetes Mellitus | 4 (5.8) |
| Intoxication | 4 (5.8) |
| Solid tumor | 4 (5.8) |
| Leukemia | 2 (2.9) |
| Nephritis | 2 (2.9) |
| Drowning | 2 (2.9) |
| Asthma | 2 (2.9) |
| Prostate hyperplasia | 2 (2.9) |
| Cerebral bleeding | 2 (2.9) |
| Electric shock | 1 (1. 5) |
| Anemia | 1 (1. 5) |
| Cystitis | 1 (1. 5) |
| Pneumonia | 1 (1. 5) |
| Encephalitis | 1 (1. 5) |
| Status thymico-lymphaticus | 1 (1. 5) |

**Figure S4. Characteristics of subjects in the Hamalainen autopsy study. A.** Age of each subject. **B.** Sex of the subjects. **C.** Causes of death of the subjects.

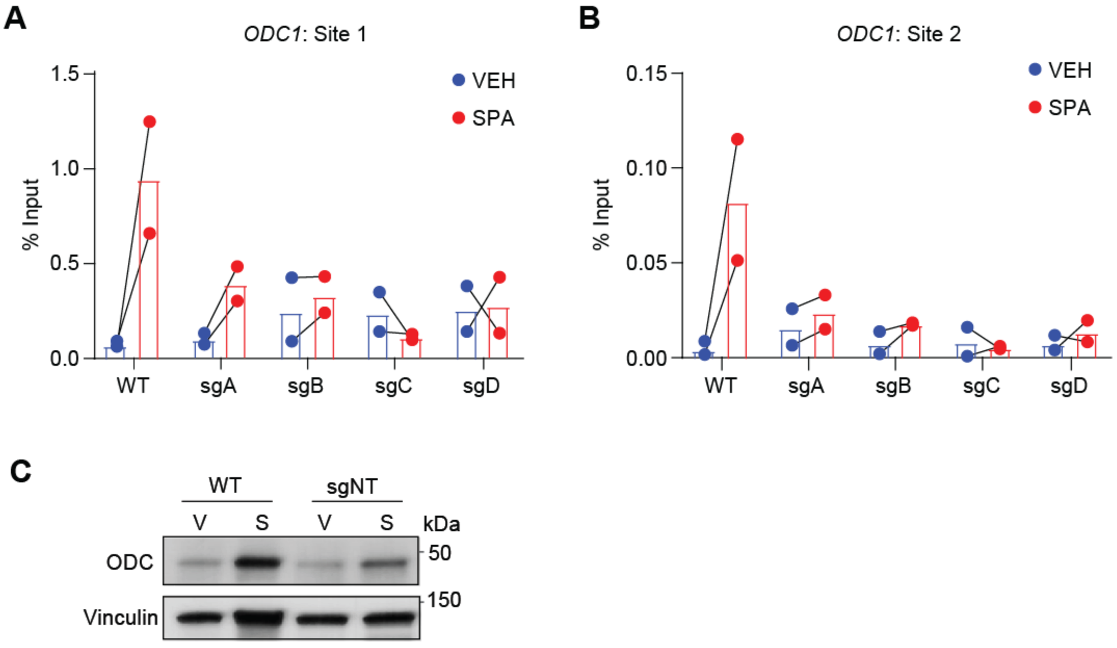


**Figure S5. dCas9-KRAB blocks AR binding upstream of *ODC1*. A-B.** AR binding at *ODC1*-site 1 (**A**) and *ODC1*-site 2 (**B**) by AR ChIP followed by RT-PCR for wild-type (WT) LNCaP cells or expressing dCas9-KRAB and sgA-D treated with vehicle control (VEH; EtOH 0.1%) or supraphysiological androgen (SPA; R1881 10nM) expressed as percent of input. N=2 independent experiments. **C.** ODC protein expression in WT LNCaP cells or cells expressing dCas9-KRAB and a non-targeting sgRNA (sgNT) treated with vehicle control (V; EtOH 0.1%) or supraphysiological androgen (S; R1881 10nM). Vinculin is used as a loading control. Representative blots of N=2 independent experiments.

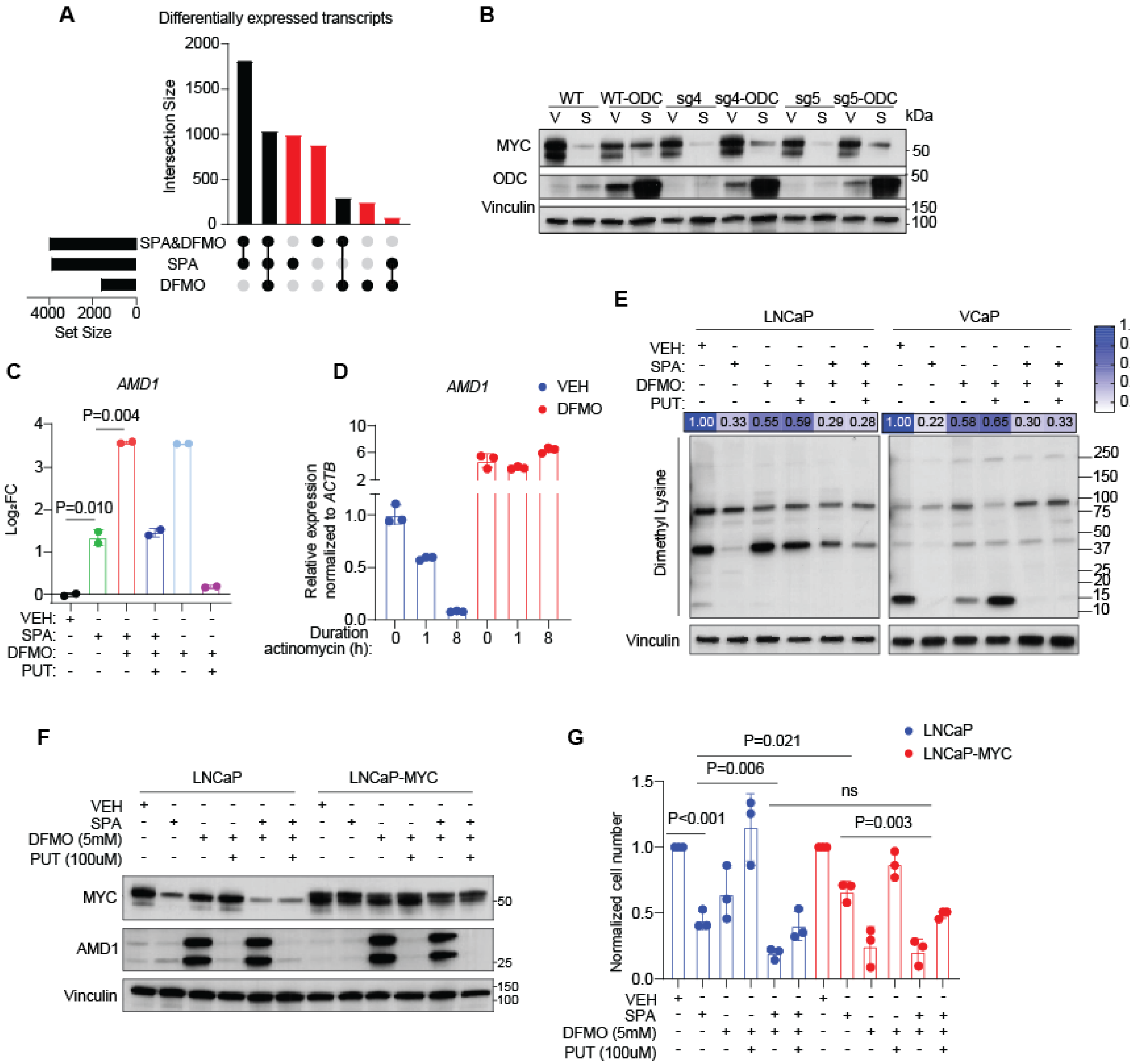


**Figure S6. Inhibition of ODC increases downregulation of MYC by SPA by depleting S-adenosylmethionine. A.** Overlap of differentially expressed transcripts occurring due to treatment as per Fig 6A. **B.** Protein expression of MYC and ODC in LNCaP cells expressing dCas9-KRAB and sgRNA-C or sgRNA-D targeting site 2 upstream of *ODC1* with and without concurrent constitutive *ODC1* cDNA expression treated with vehicle control (V; EtOH 0.1%) or supraphysiological androgen (S; R1881 10nM) for 6 days. Vinculin is used as a loading control. Representative blot of n=2 independent experiments. **C.** Change in AMD1 transcript expression in LNCaP cells treated with vehicle control (VEH; EtOH 0.1%) or combinations of supraphysiological androgen (SPA; R1881 10nM), DFMO (5mM), and putrescine (100uM) as indicated for 96 hours. **D.** Relative expression of *AMD1* transcript by RT-PCR in LNCaP cells treated with VEH or DFMO for 96 hours then actinomycin 5 ug/ml for 0, 1, or 8 hours as indicated. **E.** Abundance of proteins with dimethylated lysines in LNCaP and VCaP cells following treatment with vehicle control (VEH; EtOH 0.1%) or combinations of supraphysiological androgen (SPA; R1881 10nM), DFMO (5mM), and putrescine (100uM) as indicated for 96 hours. Vinculin is used as a loading control. Representative blots of n = 2 independent experiments. **F**. Protein expression of MYC and AMD1 in LNCaP-EV or LNCaP-MYC cells treated with vehicle control (VEH; EtOH 0.1%)

or combinations of supraphysiological androgen (SPA; R1881 10nM), DFMO (5mM), and putrescine (100uM) as indicated for 7 days. Vinculin is used as a loading control. Representative blot of n = 2 experiments. **G.** Normalized viable cell number of LNCaP-EV or LNCaP-MYC cells treated as per (A). N=3 independent experiments. P values by unpaired 2-tailed t test.

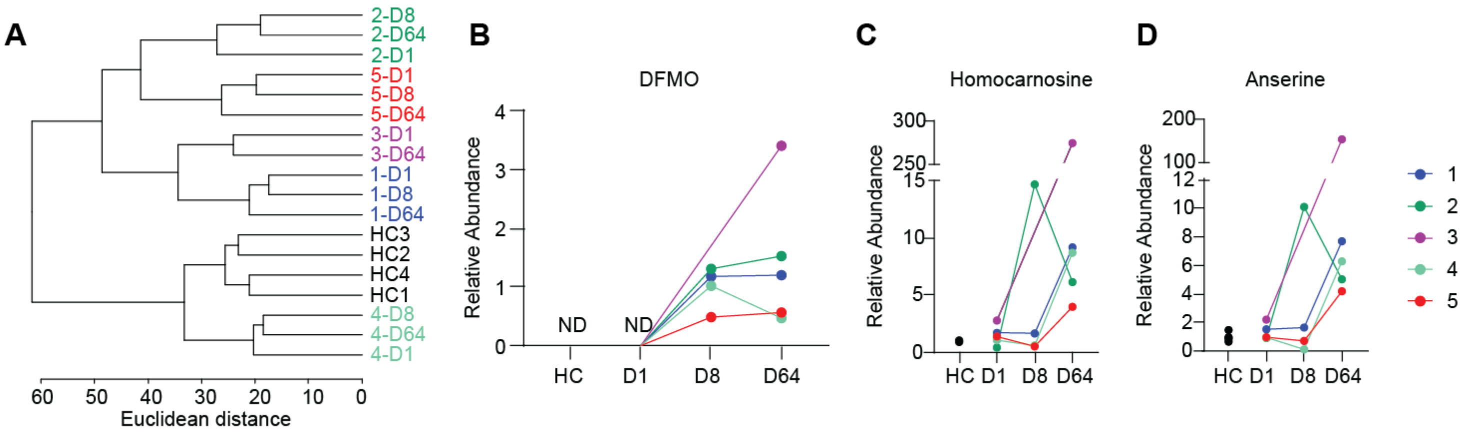


**Figure S7. Plasma metabolite analyses of trial participants A.** Euclidean clustering of fasting metabolite abundance in plasma of the first five patients on trial at indicated timepoints and 4 healthy controls (HC). **B-D.** Relative abundance of DFMO (**B**), homocarnosine (**C**), and anserine (**D**) in plasma of HC and trial participants at indicated timepoints.

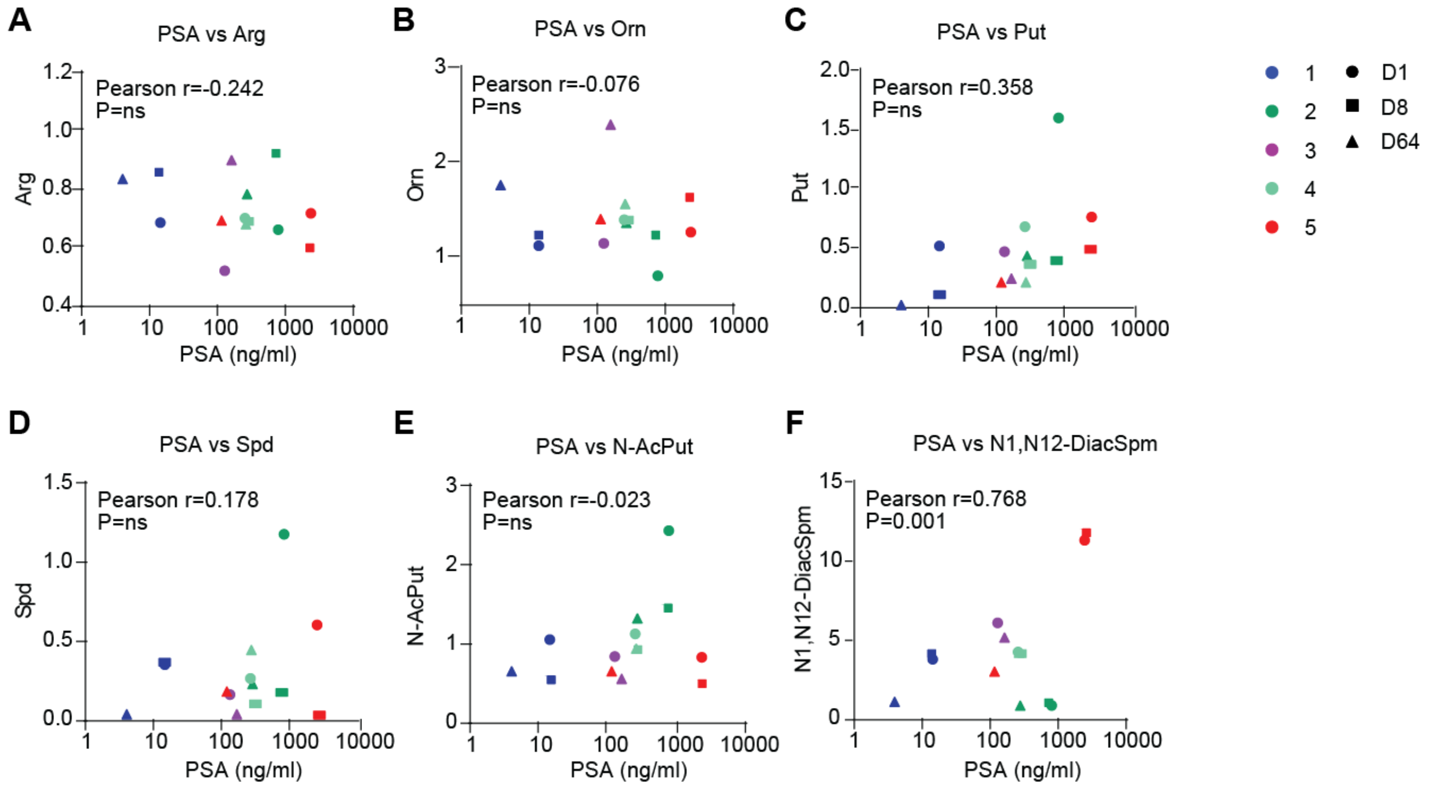


**Figure S8. Correlation of metabolites and PSA. A-F.** Correlation of plasma relative abundance of arginine (**A**), ornithine (**B**), putrescine (**C**), spermidine (**D**), N-acetylputrescine (**E**), and N1,N12-diacetylspermidine (**F**) and serum PSA. R and p values determined by Pearson's correlation calculation.

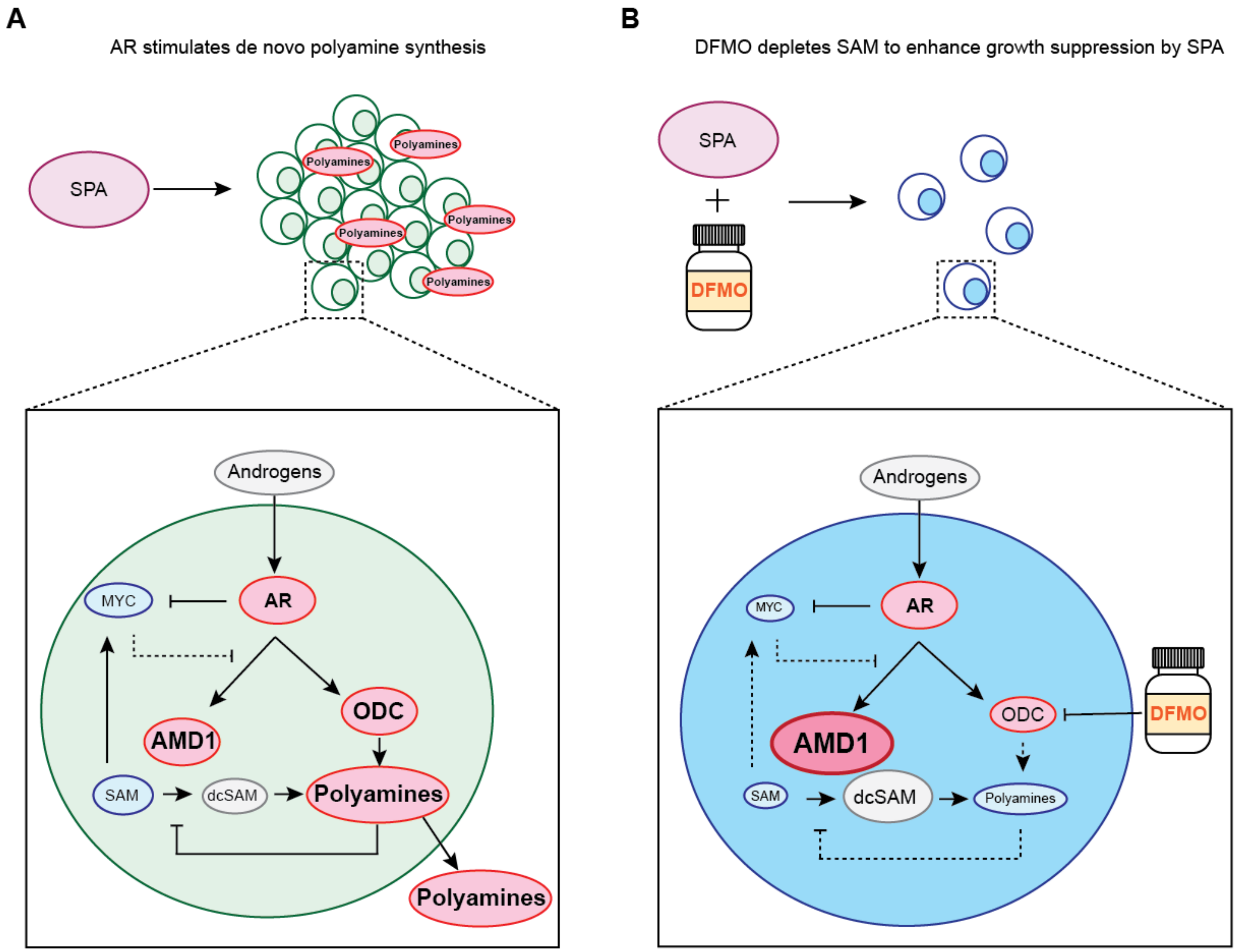


**Figure S9. Schematic summary of findings. A.** AR activation by SPA stimulates de novo synthesis of polyamines by increasing abundance of ODC and AMD1 and inhibits MYC. Decreased MYC amplifies AR-mediated induction of ODC and AMD1. Increased polyamines feedback to negatively regulate AMD1 abundance to temporize depletion of SAM and downregulation of MYC. **B.** DFMO addition to SPA inhibits ODC activity leading to depletion of polyamines and reduced negative feedback on AMD1. Enhanced AMD1 activity leads to depletion of SAM and further downregulation of MYC.
